# Empirical evaluation of the sensitivity of background incidence rate characterization for adverse events across an international observational data network

**DOI:** 10.1101/2021.06.27.21258701

**Authors:** Anna Ostropolets, Xintong Li, Rupa Makadia, Gowtham Rao, Peter R. Rijnbeek, Talita Duarte-Salles, Anthony G. Sena, Azza Shaoibi, Marc A. Suchard, Patrick B. Ryan, Daniel Prieto-Alhambra, George Hripcsak

**Affiliations:** Columbia University Medical Center, New York, NY, USA; Centre for Statistics in Medicine, NDORMS, University of Oxford, Oxford, United Kingdom; Janssen Research and Development, Titusville, NJ, USA; Department of Medical Informatics, Erasmus University Medical Center, Rotterdam, The Netherlands; Fundacio Institut Universitari per a la recerca a l’Atencio Primaria de Salut Jordi Gol i Gurina (IDIAPJGol), Barcelona, Spain; Department of Biostatistics, Fielding School of Public Health, University of California, Los Angeles, CA, USA; Department of Human Genetics, David Geffen School of Medicine at UCLA, University of California, Los Angeles, CA, USA; New York-Presbyterian Hospital, New York, NY, USA

**Author notes:** Joint first authors (Anna Ostropolets, Xintong Li). Corresponding author: George Hripcsak, Columbia University, 622 West 168th Street, PH-20, New York, NY, USA.

## Abstract

Background incidence rates are routinely used in safety studies to evaluate the association of an exposure and an outcome. Systematic research on the sensitivity of background rates to the choice of the study parameters is lacking. We used 12 electronic health record and administrative claims data sources to calculate incidence rates of 15 adverse events. We examined the influence of age, race, sex, database, time-at-risk start (anchoring) event and duration, season and year, prior observation and clean window. For binary comparisons, we calculated incidence rate ratios and performed random-effect model meta-analysis. Background rates were highly sensitive to demographic characteristics of the population, especially age, with rates varying up to a factor of 1,000 across age groups. Rates varied by up to a factor of 100 by database. Incidence rates were highly influenced by the choice of anchoring (e.g., health visit, vaccination, or arbitrary date) for the time-at-risk start, especially at short times at risk, and less influenced by secular or seasonal trends. Therefore, comparing background to observed rates requires appropriate adjustment, and results should be interpreted in the context of design choices.

**Short Figure:** 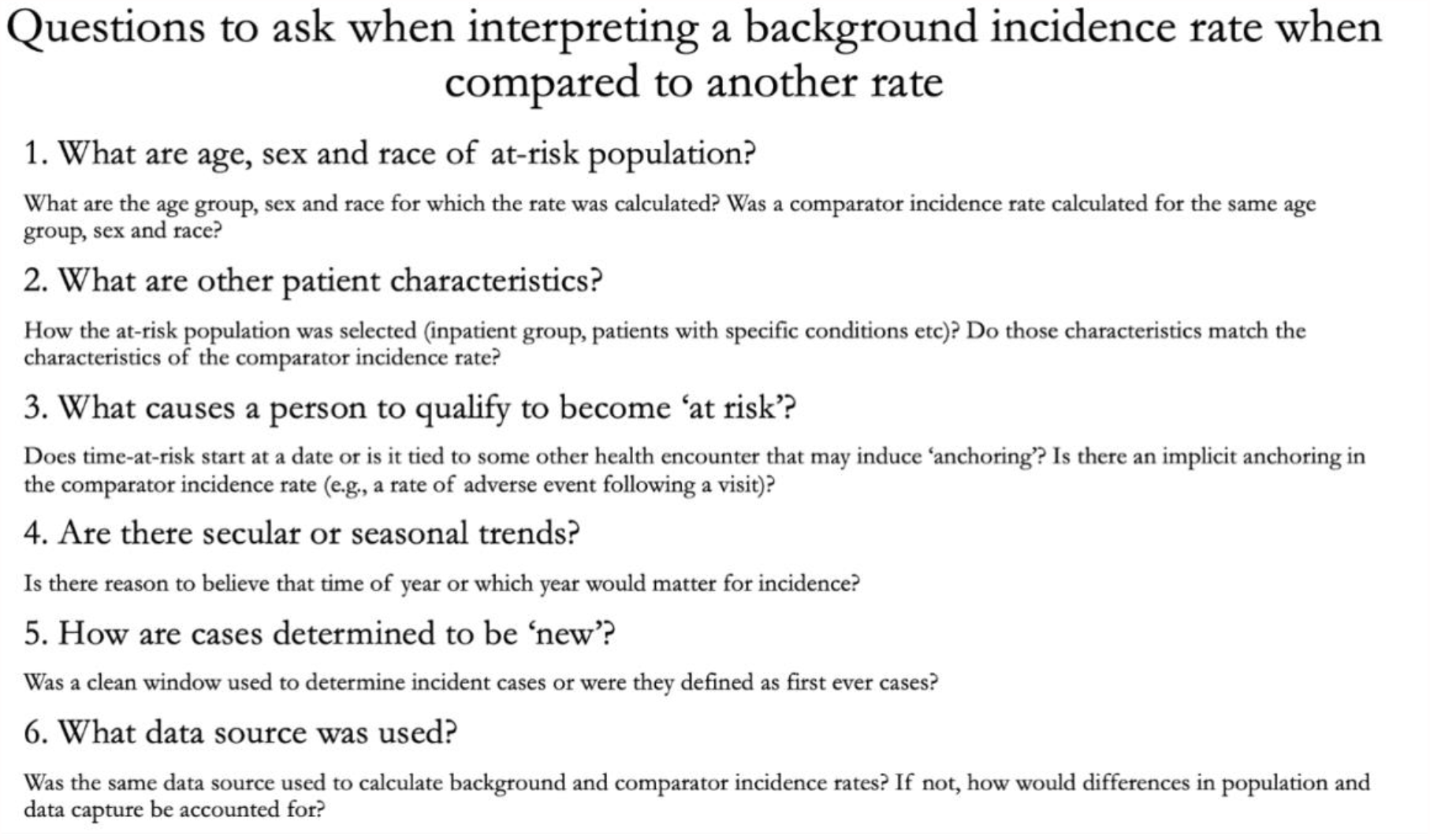

## BACKGROUND

Observational healthcare data can enable large-scale medical product safety monitoring through the identification adverse events in the data sources and application of population-level effect estimation methods to detect a possible rise in the incidence of adverse events following exposure. One approach commonly used in vaccine surveillance is to compare the observed incidence of adverse events following immunization with the background incidence in the target population (1).

As previously shown (1,2), monitoring for previously unrecognized adverse events requires accurate capture of baseline incidence rates (IRs) of events which becomes especially relevant for safety monitoring in new patient populations or mass preventative measures such as vaccination campaigns. While baseline IRs are commonly calculated in observational studies, there is little knowledge of factors influencing incidence estimation and the magnitude of such influence, which may lead to biased inference about vaccine or drug safety.

Many safety studies rely on the same data source to estimate the background incidence and the observed incidence (2). They hypothesize that the target population used to estimate the background incidence is generalizable to the patients exposed. While prospective studies allow standardized enrollment, retrospective observational studies are oftentimes performed on data sources that capture heterogeneous populations. It is unclear to what extent such populations can serve as a proxy for a counterfactual of the exposed population and whether such deviation between the comparator and that counterfactual represents a potential bias.

Patient characteristics such as age (3–6), sex (6–9), race or ethnicity (10–12) and patient location (3,9,12,13) have been shown to have an impact on the IRs. For example, the studies reported up to a 10-fold difference in IRs of adverse events in different age groups (1), up to a 20-fold difference in IRs across different data sources (6) and a 7-fold age-adjusted difference in IRs in different ethnic populations (11). There is also limited research on other patient characteristics that may influence baseline IRs, such as patient primary healthcare institution (4,6,14,15) or prior observation time (16). Nevertheless, the influence of demographics has not been studied systematically.

There is also a lack of research on quantifying the impact of time-at-risk (TAR) window choice on baseline IRs. While this choice is usually based on the pharmacokinetics and pharmacodynamics of the drug, there is no clear algorithm to choose such a window. Proposed approaches include using a set of pre-specified windows for all outcomes (1) or varying time-at-risk windows to empirically select an appropriate one (17). Other studies showed temporal trends (9,18) in IRs, which could be attributed to changes in the underlying patient population, improved diagnosis of outcomes, changes in data capture or other factors.

Another gap in research is related to the starting point used to estimate baseline IRs. Most of the studies use an arbitrary calendar date for time-at-risk start, which can be the date patients satisfy the inclusion criteria or start of the year for annual IRs. On the other hand, anchoring time-at-risk interval on a healthcare encounter may be associated with observing more adverse events due to the impact of administered drugs or detection bias.

Indeed, there is no common framework to assess baseline IRs in drug safety and effectiveness studies, which results in high heterogeneity of IRs in most of the meta-analyses of IRs (19–22). Even if the above-mentioned factors were taken into consideration, variation methods to standardize baseline IRs, including stratification by age or sex, adjusting for age of the US population, or simply reported multiple unadjusted IRs, may disable comparison of such estimates.

With the ongoing COVID-19 vaccination campaign, several regulatory bodies have published the protocols to assess background rates, which differ in data sources used, requirements for prior observation periods, anchoring date, clean window and outcome definitions (23,24). Recent papers on estimating background rates of adverse events of interest for COVID-19 vaccine also used heterogeneous definitions and settings (25–27). Such discrepancies may result in producing different incidence rates and obscure their interpretation. In this paper, we provide an analysis of parameters influencing background rate estimation and discuss implications for interpreting incidence rates.

## METHODS

Our primary research question was: “How does the selection of analysis parameter choices (such as target population, anchoring event, time-at-risk, and data source) influence baseline incidence rate estimation?” To address it, we identified the set of choices related to each part of the incidence rate estimation (Figure 1) and specified experiments to estimate the sensitivity to those parameter choices.

**Figure 1.**
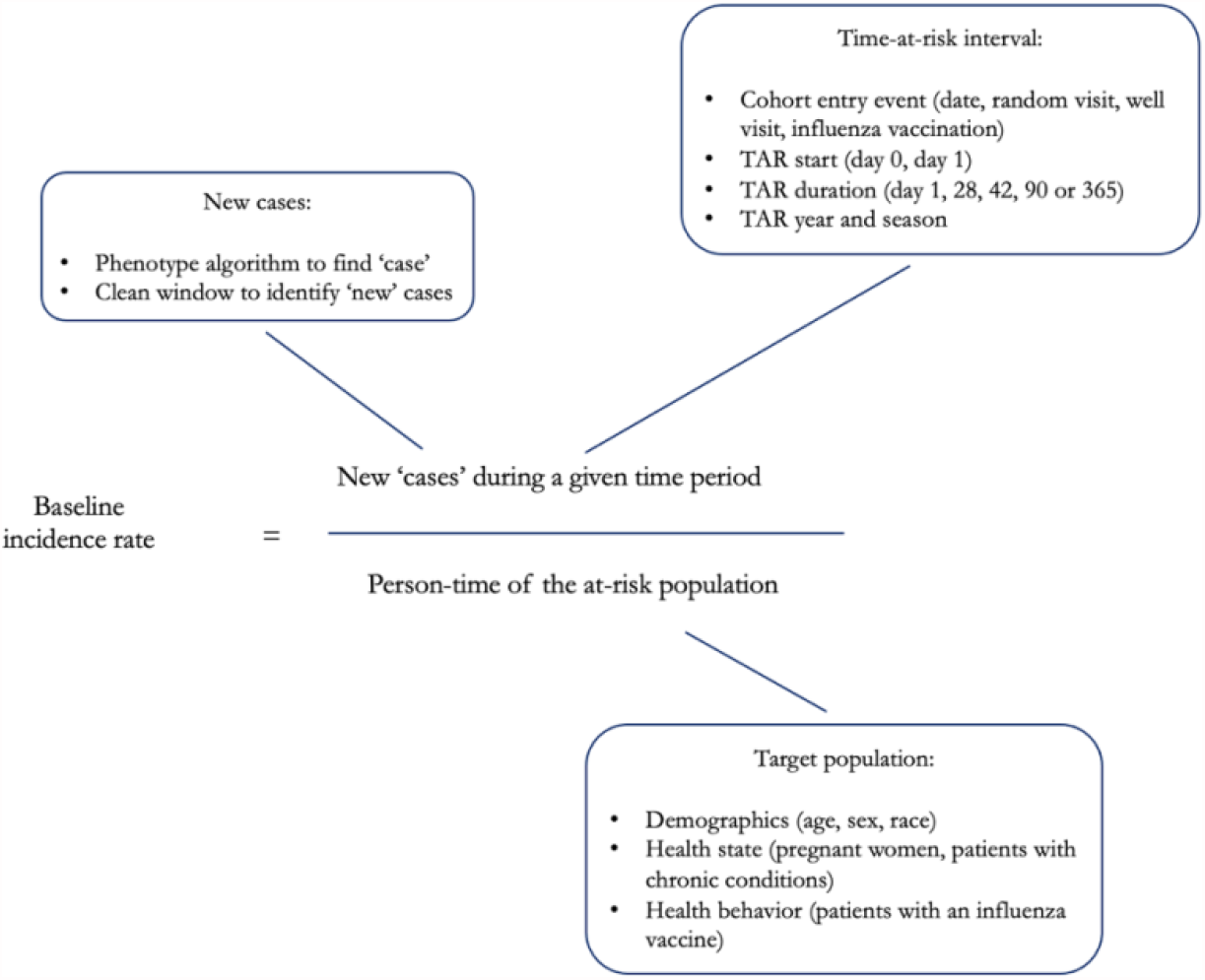
Baseline incidence rate calculation and its elements.

### Baseline incidence rate estimation analysis parameter choices

#### Data sources

We conducted the experiment on 12 data sources (Table S1), including sources with different data source provenance (administrative claims data, electronic health record data), origin (the US, Australia, Germany, France, Japan, the UK), and representing different populations (privately insured employed patients in IBM MarketScan Commercial Claims (CCAE) or patients with limited income in IBM MarketScan Multi-state Medicaid (MDCD)).

#### Phenotype creation and evaluation

We used the outcomes outlined in the “Background Rates of Adverse Events of Special Interest for COVID-19 Vaccine Safety Monitoring” protocol published by Food and Drug Administration Center for Biologics Evaluation and Research (23), which included Guillain-Barré syndrome, facial nerve (Bell’s) palsy, anaphylaxis, encephalomyelitis, narcolepsy, appendicitis, non-hemorrhagic stroke, hemorrhagic stroke, acute myocardial infarction, myocarditis and pericarditis, deep vein thrombosis, pulmonary embolism, disseminated intravascular coagulation, immune thrombocytopenia and transverse myelitis. For each outcome, we created a phenotyping algorithm to identify patients with a given condition using available structured data. After defining an initial set of patients, we explored patient characteristics (demographic information, co-morbidities, procedures and drug histories). Algorithms were iteratively defined to ensure comprehensive capture and reviewed by the team of informaticists, epidemiologists and clinicians. All phenotype algorithm definitions, including associated code lists, are provided in Table S2 and are publicly available at atlas.ohdsi.org.

We did not examine phenotypes requiring inpatient encounter on the outpatient data sources (IQVIA Australia, IQVIA Germany, IQVIA France, CPRD GOLD, ICPI Netherlands). We also excluded the phenotypes that did not yield patients on given data sources, as well as age strata less than 55 years for MDCR. Results for transverse myelitis in JMDC and narcolepsy in Optum EHR were removed due to failed cohort diagnostics.

#### Target Population

We studied three components of target population: inclusion criteria specifying which patients enter the cohort, index date at which patients enter the target cohort and patient demographics (stratification on age groups, sex and race).

#### Patient cohort definition

The base population was the patients observed in the database at any time during 2017-2019 with at least 365 days of prior observation. We also selected several subsets of the target population based on health state and behavior (Figure 1).

For patients with a well visit, the latter was defined as a healthcare encounter associated with CPT4 codes representing well visits. A chronic condition visit was defined as a healthcare encounter with at least one condition diagnostic code associated with a higher risk of complications as defined by CDC (Table S2). Pregnancy episodes were constructed using a published algorithm (28).

#### Cohort entry (anchoring)

We studied several entry date anchors:

- An arbitrary calendar date, which was set to January 1st for overall and annual IR calculation and a start of period (January 1^st^, April 1^st^, July 1^st^, October 1^st^) for seasonal incidence calculation
- First healthcare encounter (inpatient, outpatient or emergency department visit)
- First well visit
- Influenza vaccination

#### Population subgroups

For each target population, we stratified on age (0–5, 6–17, 18–35, 36–55, 56–64, 65–74, 74–85, > 85), sex (male, female) and race (White, Black).

#### Time at risk

For each outcome, we compared selected five time-at-risk windows relative to target cohort entry date (0 – 1 days, 1 – 28 days, 1 – 42 days, 1 – 90 days and 1 – 365 days) (23).

### Sensitivity experiment

We performed calculations for each combination of outcome, target population and time-at-risk. We calculated incidence rate as the ratio of the number of cases to the total person-time the population was at risk (from cohort start date to the end of time-at-risk period, occurrence of an outcome or loss to follow-up whichever comes first).

Annual IRs were calculated for 2017, 2018, 2019 and 2020. Seasonal IRs were computed across four seasons in 2017 – 2020 (1/1–3/31, 4/1–6/30, 7/1–9/30, 10/1–12/31 in each year). We also performed comparison of IRs estimated for a period of COVID-19 pandemic (4/1/2020–9/31/2020), which was compared to the same period in 2019.

To make comparisons between the incidence rates observed under different analysis settings, incidence rate ratios (IRR) were computed, holding all parameters constant except for the target parameter of interest. Comparisons using IRR included: male versus female patients, White versus Black patients, no ‘at risk’ comorbid condition versus >=1 ‘at risk’ comorbid condition, outcome-specific clean window versus no prior events as well as comparisons of different years and seasons. For all incidence rate ratios, we conducted random-effects model meta-analyses to generate age-adjusted and unadjusted pooled IRRs and 95% confidence intervals across data sources using R package metafor version 2.4 (29). Heterogeneity was assessed using the I^2^ index (30). Detailed description of analysis parameters for each experiment and detailed results can be accessed on Github study page (31).

## RESULTS

As expected, the incidence rates of the outcomes displayed a very wide range. When calculated for all age groups, target populations and anchoring events, IRs of outcomes showed more than 100,000-fold differences (Figure 2). Population-wide IRs (without age-sex stratification) varied from 0.0001 - 1.74 per 100 patients for disseminated intravascular coagulation to 0.01 - 99.2 for non-hemorrhagic stroke.

**Figure 2.**
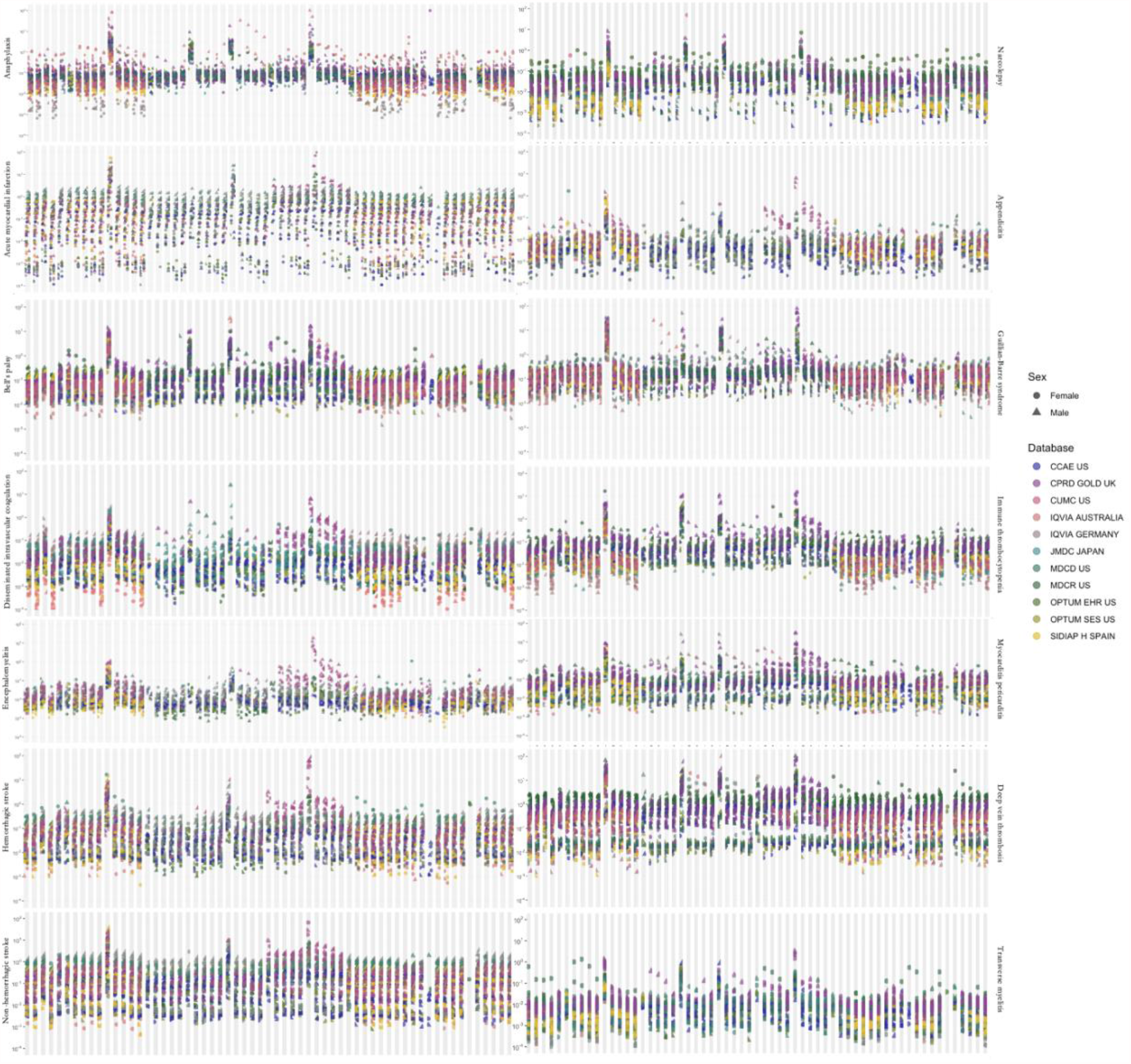
Estimated incidence rates for adverse events of interest across all a) target populations, b) time-at-risk intervals and c) age groups. A dot represents one incidence rate estimate.

Age was the main contributor to the heterogeneity showed in Figure 2, with rates varying by up to a factor of 1,000 across age groups for an outcome within one database. The effect of age was observed consistently across all data sources and outcomes, which highlights the extreme sensitivity of the incidence rate estimation to the age distribution of the measured population.

We looked at the effect of sex on incidence rate, quantifying the IRR of incidence rates in males compared to females (Table S3 and Figure S1). Most outcomes revealed a statistically significant difference, with the direction of the difference matching the literature: transverse myelitis (pooled IRR 0.8 (95% CI 0.7-0.9)) was more common in females, myocarditis and pericarditis (IRR 1.6 (95% CI 1.5-1.8)), appendicitis (IRR 1.1 (95% CI 1.1-1.2)), acute myocardial infarction (IRR 2.2 (95% CI 1.9-2.5)) and strokes (IRR 1.3 (95% CI 1.2-1.4) and IRR 1.4 (95% CI 1.3-1.5)) were more common in males.

For most of the conditions, race did not have a substantial effect on incidence rates (Table S3 and Figure S2). Disseminated intravascular coagulation, myocarditis, non-hemorrhagic stroke and pulmonary embolism were more common in Black patients and appendicitis was more common in White patients.

Figure 3 also shows the variation in incidence rates between databases even for the same outcome and age group. Differences of a factor of 10 were common, especially for rare disorders like disseminated intravascular coagulation, narcolepsy, immune thrombocytopenia and transverse myelitis. Generally, these disorders had higher incidence in the non-US data sources compared to the US data sources. Notably, disseminated intravascular coagulation higher incidence in Japan. We found that all of age-sex population strata showed at least 40% heterogeneity by I^2^ in strata- and outcome-specific meta-analyses.

**Figure 3.**
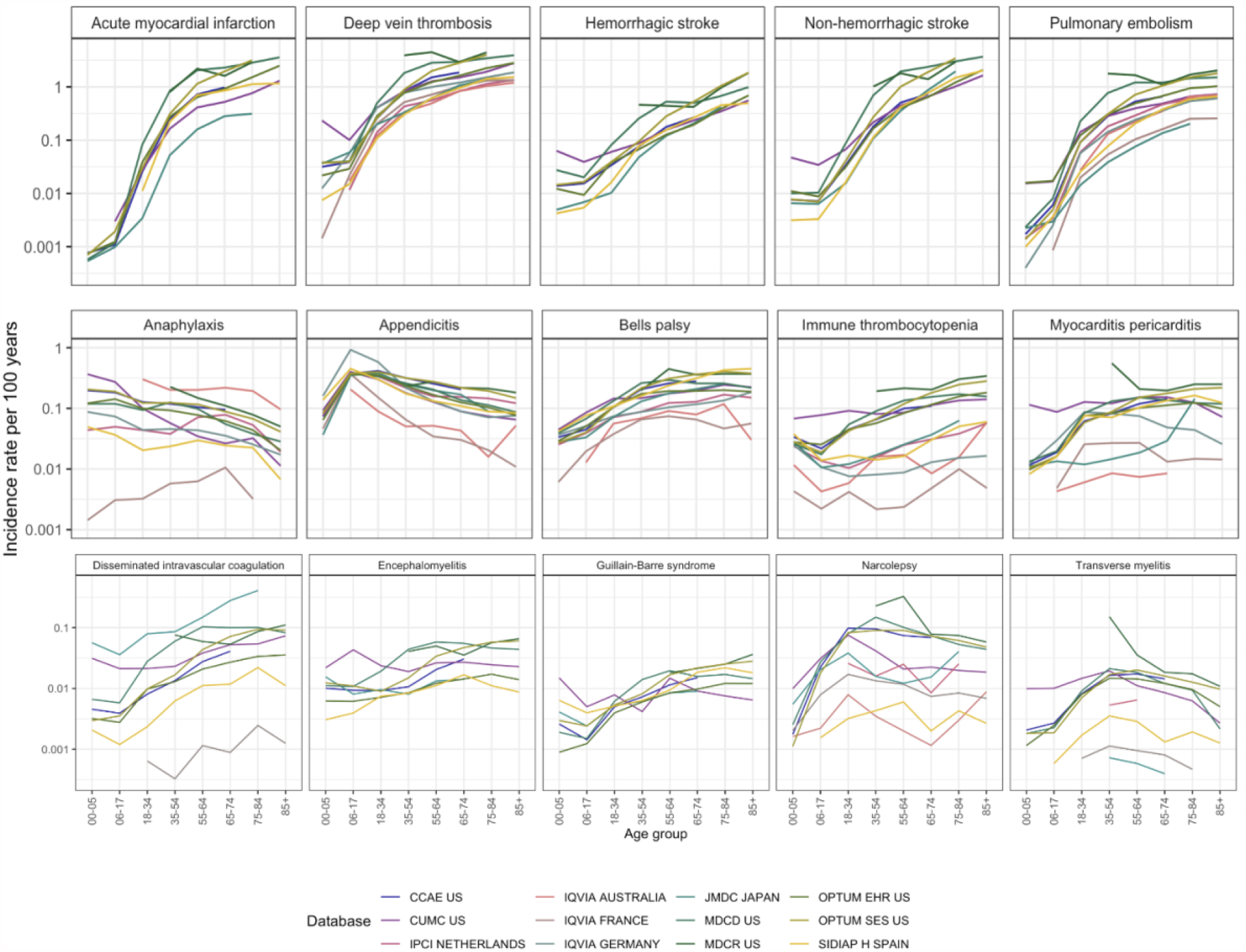
Incidence rates in age groups in 2017 – 2019 in patients entering on January 1 with a 365-day time at risk and 365 days of pre-entry observation period. Outcomes were arranged by maximum incidence per age stratum from the most common to the least common.

When adjusted for age, anchoring was the second-largest effect we found, where the IRR of anchoring on a visit versus anchoring on January 1^st^ for a short time-at-risk (0-1 day) was associated with up to a 100-fold increase in incidence (pooled IRR 26.8 (95% CI 21.9-32.8), IRR for anaphylaxis 47.6 (95% CI 32.8 – 69.1)). The effect was attenuated for longer times at risk (Figure 4). For a time at risk of 28 days, the IRR was 1.4 (95% CI 1.3-1.5) and smaller with longer time-at-risk intervals (Table S4).

**Figure 4.**
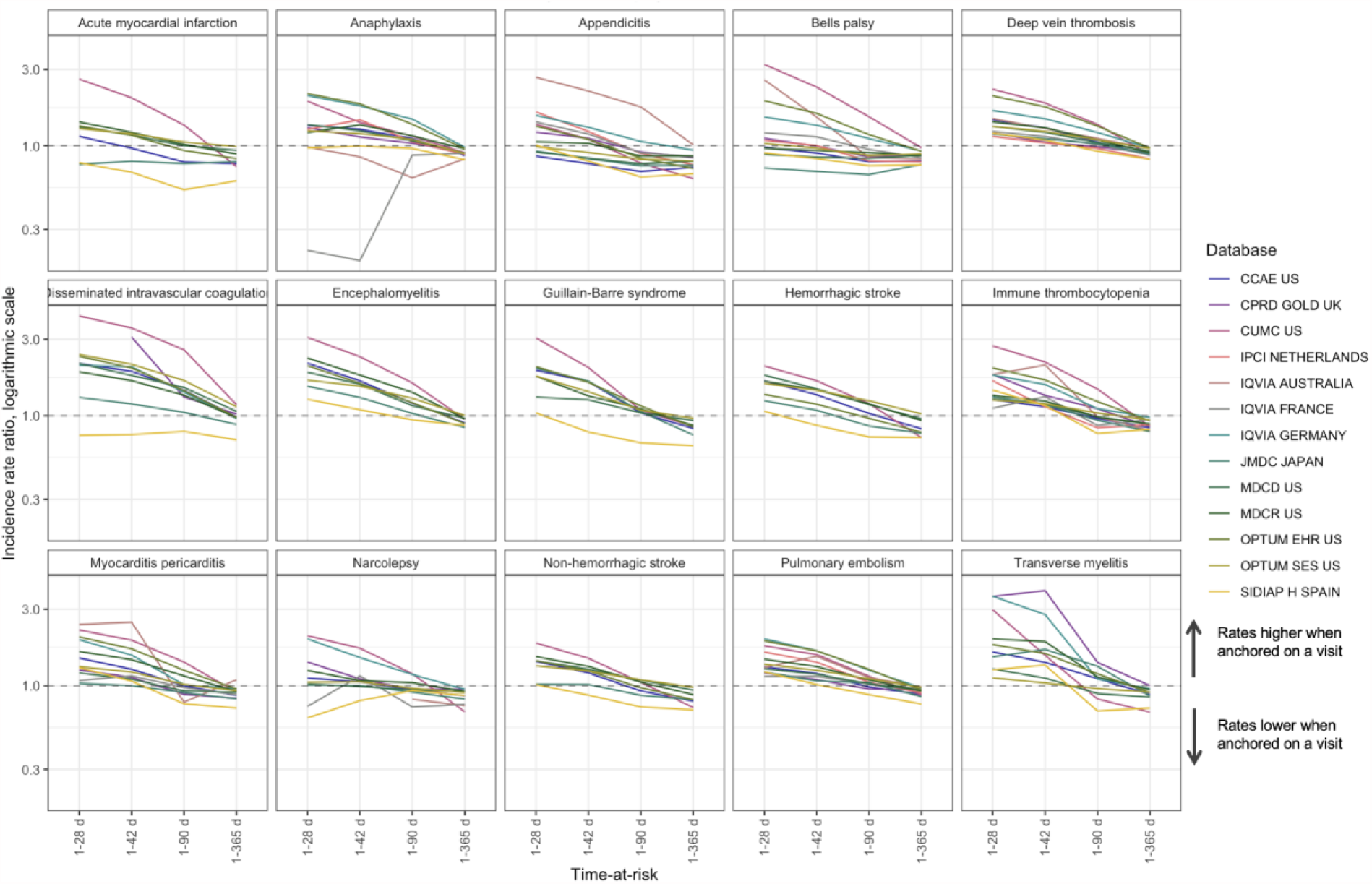
Comparison of anchoring on a random visit versus anchoring on January 1^st^ in patients with a visit in the next year for time-at-risk 1-28 days, 1-42 days, 1-90 days and 1-365 days, incidence rate ratio.

Additionally, we found that when anchoring on a visit, the incidence rates for 1-365 day time-at-risk were lower than in the group of patients with a visit in the next year anchored on January 1^st^. This may be explained by the fact that anchoring on a visit excludes the day of the visit from time-at-risk, while time-at-risk for anchoring on January 1^st^ includes the days of subsequent visits. Including day 0 in time-at-risk mitigates this difference (Table S4).

We observed similar trends for anchoring on a well visit or an influenza vaccination with the pooled incidence rate ratios 1.21 (95% CI 1.11-1.31) and 1.17 (95% CI 1.11-1.22) respectively (Tables S5 and S6, Figures S4 and S5). Some of the conditions were influenced by anchoring on any type of visit (for example, disseminated intravascular coagulation and acute myocardial infarction). Notably, incidence of Guillain-Barre syndrome was significantly increased when anchoring on an influenza vaccination and was less influenced by anchoring on a well visit or a random visit.

Time at risk influenced incidence only when we anchored on an event. When anchoring on January 1^st^, comparing the time at risk for 1 day versus 365 days showed consistently little effect across all outcomes with the pooled IRR across databases and outcomes of 1.0 (95% CI 0.93-1.08).

We observed seasonal trends for anaphylaxis, appendicitis, acute myocardial infarction, strokes and Guillain-Barre syndrome (Figure S6 and Table S7). We also found a decrease in IRs in some of the data sources in 2020 compared to 2019-2017, which may be attributable to the changes in healthcare delivery during COVID-19 pandemic (Figure S7 and Table S8). Additionally, we analyzed incidence rates of outcomes in two subgroups of interest for COVID-19 vaccine: pregnant women and patients with chronic conditions. The latter group had significantly higher incidence rates of all outcomes when compared to the group of patients with no chronic conditions (pooled IRR 2.16, 95% CI 1.91 - 2.44). Alternatively, when looking at the patients with prior influenza vaccination (as a proxy for patients willing to receive a vaccine), we found that patients with prior influenza vaccination had higher incidence of all outcomes compared to the general population (pooled IRR 1.41, 95% CI 1.30 - 1.54, Table S9 and Figure S8).

In this study, we defined incident cases as those that occurred for the first time in a given window defined by the FDA BEST protocol. When comparing an alternative approach for defining incident cases – cases that occurred for the first time in the patient history – we found that using all patient history to define incident cases produces consistently smaller incidence rates for all outcomes with the pooled IRR of 0.83 (95% CI 0.79-0.87). Notably, IRRs for narcolepsy and Guillain-Barre syndrome were significantly smaller (IRR 0.69 (95% CI 0.65-0.74) and IRR 0.59 (95% CI 0.48-0.71) respectively, Table S10 and Figure S9).

This observation was supported by modestly lower incidence when requiring patients to have prior observation in the database to ensure data capture with the pooled IRR of 0.94 (95% CI 0.9 – 0.99). While this trend was not observed for all outcomes, narcolepsy, Guillain-Barre syndrome and myocarditis yet again were greatly impacted by a requirement of prior observation (Table S10 and Figure S9).

## DISCUSSION

In this study, we observed wide variation of incidence rates depending on the study parameters. The population characteristics such as age and sex had the largest impact with incidence rates varying up to a factor of 1,000 across age groups. Even after adjusting for age and sex, the study showed signs of residual bias due to the other parameters. Anchoring, which was not previously considered in incidence rate studies, appeared to be the second most important parameter. Anchoring on any type of healthcare encounter yielded higher incidence when compared to anchoring on a random date, especially for the short time-at-risk. Duration of time-at-risk interval mattered a lot with the short time-at-risk intervals associated with a higher incidence rates for all conditions. When incident cases were defined using all patient history as opposed to pre-defined clean windows, observed incidence rates were higher. In the context of safety studies, some of the abovementioned factors can be adjusted for in the analysis, while others have to be accounted for in study design.

Post-marketing safety surveillance aims at monitoring previously unrecognized serious events following medical product exposure. Active surveillance is especially relevant in the context of COVID-19 vaccination, where large populations are being exposed in a relatively short duration, heightening the need to detect a possible rise in the incidence of adverse events in a timely manner. As observed rates of events are compared to the background incidence in a population assessing causality requires accurate identification of background rates (1,2). Background rates are sensitive to each of the components of IRs estimation.

### Population at risk: age, sex, race

In order to appropriately use background incidence in the context for interpreting the observed rate of adverse events in a defined exposed population, it is important to consider the extent to which the target population that is used to estimate the background incidence is generalizable to the exposed individuals. In any observed vs. expected comparison, the comparator serves as a proxy for a counterfactual of the exposed population — what would have happened to those same individuals had they not been exposed — and any deviation between the comparator and that counterfactual represents a potential bias. It is, therefore, important, to access target population characteristics and estimate their influence on background rates.

Age is one of the key characteristics of the target population that was previously shown to influence incidence rates (3,5,6,32). Our study shows the extreme size of the age effect on the incidence of all outcomes across all data sources (23). Sex was also observed to influence incidence rates with sex trends matching previous literature (5,33–36). One has to perform age and sex adjustment stratification when comparing background and observed incidence rates. Similarly, race contributed to differences in incidence rates, especially for cardiovascular conditions.

### Database effects

Large effect of data source choice is likely a combination of actual population differences—age, sex, race, acuity, differences in genetics and environmental exposure—as well as differences in measurement, such as collection via administrative claims versus electronic health records. The differences suggest that, where possible, background rates should be calculated in the database where the surveillance will be done. Where this is not possible, a broad range of databases should be used and, based on a random-effects meta-analysis, prediction intervals should be calculated for the incidence rates. Prediction intervals will unfortunately be wide, but they will reflect the true uncertainty of the incidence rates of a surveillance database in which baseline rates cannot be measured (e.g., because it is a new data source). We demonstrate our prediction intervals in Table S12.

### Large effect of anchoring on health encounters

Despite age and sex adjustment and even calculating rates in the same database, other factors have to be accounted for when interpreting incidence rates. Anchoring, or cohort entry criteria, was the second parameter that influenced incidence rates, especially for the short time-at-risk windows. As most of the random visits are associated with patients’ conditions, having a visit was associated with an increase in existing and new condition capture as well as a potential increase in adverse events following interventions or treatments administered or prescribed during the visit. This effect was not quantified before and, surprisingly, was also present for anchoring on a well visit, which had been hypothesized to indicate visits not associated with conditions or symptoms.

When studying background incidence in the context of COVID-19 vaccination (in cohort or self-controlled studies), estimation of incidence rates of events following vaccination is anchored on the date of vaccination. To appropriately compare it to the background rates, one has to make an assumption of the type of encounter that represents the vaccination best. For example, in a wide population that receives the vaccine based on availability, a random date may be a good approximation for the date of vaccination. On the other hand, vaccination date in patients receiving vaccine upon hospital discharge or in nursing homes may represent a strong anchor with the effect similar to or even greater than anchoring on a random visit.

Influenza vaccination may serve as another proxy for COVID-19 vaccination, in terms of defining an anchoring event. While anchoring on vaccination was associated with a slight increase in incidence, the latter for disproportionally increased for Guillain-Barre syndrome, which may reflect a causal relationship (34). Nevertheless, the population that receives an influenza vaccine in healthcare institutions may be different from those who receive it in drug stores (38). It may explain why we observed higher incidence of conditions in patients with prior influence vaccine as vaccine in this case may be indicative of co-morbid conditions.

### Muted seasonal effect and small annual increase

While previous research emphasized the influence of season on IRs (40), we observed that seasons did not affect the incidence of disorders much. Myocarditis and myocardial infarction were observed at a slightly higher rate in winter and narcolepsy – at a slightly lower rate, which agreed with the literature (3,41,42). Notably, anaphylaxis was greatly influenced by the season and was more common in summer, which was previously attributed to serum, venom and food but not to medication triggers (43). Temporal trends were moderate: incidence rates slightly increase from 2017 to 2019, which may correspond to better diagnosis or changes in coding practices. That agrees with the findings in the literature for encephalomyelitis, hemorrhagic stroke, anaphylaxis, narcolepsy, Bell’s palsy (6,44–48).

### Incident cases

Patient phenotyping plays a crucial role in the incidence rate estimation. As previously shown, different phenotype definitions can yield up to 20-fold difference in incidence rates even when the studies was conducted on the same data source (49–52).

While examining different phenotypes was not the focus of this study, we found that using a clean window for excluding patients with prior disorders as opposed to using all patient history yields lower Irs for all outcomes. The observed difference in the incidence of chronic conditions or conditions that are likely to occur once (such as appendicitis) reflects potential index event misclassification. It is possible, that such patients are captured later in the course of the disease, which requires thoughtful examination of the patient history to determine the true condition start date.

Using a requirement of prior observation ensures that patients were actively observed in the data source. In this study, we found that such a requirement did not produce a difference in IRs when compared to the broad population. On the other hand, it potentially reduces index event misclassification as more information about the patient is captured.

## LIMITATIONS

Due to observational nature of the study, the data sources may not have complete capture of patient conditions. While our phenotype algorithms may be subject to measurement error, such error in unlikely to be differential. As the goal of the study was not to establish causality but to estimate sensitivity of incidence rates, phenotype measurement error or partial data capture should not influence the results of the study. As race is available only in some data sources, our findings regarding race influence may not be generalizable to other data sources or populations.

## CONCLUSIONS

Accurate estimation of background rates is essential for conducting safety or effectiveness studies. Background incidence rates are highly sensitive to demographic characteristics of population, so estimation requires age, sex and race adjustments. Even when adjusted for these factors, incidence rates are highly influenced by the choice of the time-at-risk start date or event. When comparing background rates to estimated incidence rates, one has to examine if the choice of anchoring is compatible between groups. If anchored, short time-at-risk intervals are associated with higher incidence, so the choice of time-at-risk requires thoughtful analysis. Similarly, the choice of clean window for defining incidence cases results in different incident rates. Finally, the choice of year and season may influence rates, albeit the influence is not prominent compared to the other factors.

## Data Availability

Patient-level data cannot be shared without approval from data custodians due to local information governance and data protection regulations. Aggregated data, analytical code, and detailed definitions of algorithms for identifying the events are available in a GitHub repository (https://github.com/ohdsi-studies/Covid19VaccineAesiIncidenceCharacterization).

## Ethical approval

The protocol for this research was approved by the Columbia University Institutional Review Board (AAAO7805), the Independent Scientific Advisory Committee (ISAC) for MHRA Database Research (20_000211), the IDIAPJGol Clinical Research Ethics Committee (project code: 21/007-PCV), the IPCI governance board (application number 3/2021).

