## Supplementary Materials for "Empirical evaluation of the sensitivity of background incidence rate characterization for adverse events across an international observational data network"

**Supplements**

**Table S1**. Data sources

| Data source | Description |
|---|---|
| IQVIA Australia Electronic Medical Records (Australia EMR) | Anonymized patient records of more than 6 million patients in Australia collected from Patient Management software used by GPs during an office visit to document patients' clinical records. |
| IQVIA Longitudinal Patient Data France (LPD France) | Anonymized patient records of 7.8 million patients in France collected from Patient Management software used by GPs and select specialists during an office visit to document patients' clinical records |
| IQVIA Disease Analyser Germany (DA Germany) | IQVIA DA Germany is collected from extracts of patient management software used by GPs and specialists practicing in ambulatory care settings. Data coverage includes more than 34M distinct person records out of at total population of 80M (42.5%) in the country and collected from 2,734 providers. Dates of service include from 1992 through March 2020. |
| Japan Medical Data Center (JMDC) | Japan Medical Data Center (JMDC) database consists of data from 60 society-managed health insurance plans covering workers aged 18 to 65 and their dependents (children younger than 18years old and elderly people older than 65 years old). JMDC data includes membership status of the insured people and claims data provided by insurers under contract (e.g. patient-level demographic information, inpatient and outpatient data inclusive of diagnosis and procedures, and prescriptions as dispensed claims information). |
| Optum© de-identified Electronic Health Record Dataset (Optum EHR) | Optum© de-identified Electronic Health Record Dataset is derived from dozens of healthcare provider organizations in the United States (that include more than 700 hospitals and 7,000 Clinics treating more than 103 million patients) receiving care in the United States. The medical record data includes clinical information, inclusive of prescriptions as prescribed and administered, lab results, vital signs, body measurements, diagnoses, procedures, and information derived from clinical Notes using Natural Language Processing (NLP). |
| Optum® De-Identified Clinformatics® Data Mart Database – Socio-Economic Status (Optum SES) | Optum© De-Identified Clinformatics® Data Mart Database (Optum Insight, Eden Prairie, MN) is an adjudicated administrative health claims database for members with private health insurance, who are fully insured in commercial plans or in administrative services only (ASOs), Legacy Medicare Choice Lives (prior to January 2006), and Medicare Advantage (Medicare Advantage Prescription Drug coverage starting January 2006). The population is primarily representative of US commercial claims patients (0-65 years old) with some Medicare (65+ years old) however ages are capped at 90 years. It includes data captured from administrative claims processed from inpatient and outpatient medical services and prescriptions as dispensed, as well as results for outpatient lab tests processed by large national lab vendors who participate in data exchange with Optum. Optum SES provides |

| | |
|---|---|
| | socio-economic status for members with both medical and pharmacy coverage and location information for patients at the US Census Division level. |
| IBM MarketScan Medicare Supplemental and Coordination of Benefits Database (MDCR) | IBM MarketScan Medicare Supplemental and Coordination of Benefits Database (MDCR) represents health services of retirees in the United States with primary or Medicare supplemental coverage through privately insured fee-for-service, point-of-service, or capitated health plans. These data include adjudicated health insurance claims (e.g. inpatient, outpatient, and outpatient pharmacy).Additionally, it captures laboratory tests for a subset of the covered lives. |
| IBM MarketScan Multi-State Medicaid Database (MDCD) | IBM MarketScan Multi-State Medicaid Database (MDCD) contains adjudicated US health insurance claims for Medicaid enrollees from multiple states and includes hospital discharge diagnoses,outpatient diagnoses and procedures, and outpatient pharmacy claims as well as ethnicity and Medicare eligibility. Members maintain their same identifier even if they leave the system for a brief period; however the dataset lacks lab data. |
| IBM MarketScan Commercial Claims and Encounters Database (CCAE) | IBM MarketScan Commercial Claims and Encounters Database (CCAE) is a US employer-based private-payer administrative claims database. The data include adjudicated health insurance claims (e.g. inpatient, outpatient, and outpatient pharmacy) as well as enrollment data from large employers and health plans who provide private healthcare coverage to employees, their spouses, and dependents. Additionally, it captures laboratory tests for a subset of the covered lives. This administrative claims database includes a variety of fee-for-service, preferred provider organizations, and capitated health plans. |
| Clinical Practice Research Datalink (CPRD) | The Clinical Practice Research Datalink (CPRD) is a governmental, not-for-profit research service, jointly funded by the NHS National Institute for Health Research (NIHR) and the Medicines and Healthcare products Regulatory Agency (MHRA), a part of the Department of Health, United Kingdom (UK). CPRD consists of data collected from UK primary care for all ages. This includes conditions, observations, measurements, and procedures that the general practitioner is made aware of in additional to any prescriptions as prescribed by the general practitioner. In addition to primary care, there are also linked secondary care records for a small number of people. The major data elements contained within this database are outpatient prescriptions given by the general practitioner (coded with Multilex codes) and outpatient clinical, referral, immunization or test events that the general practitioner knows about (coded in Read or ICD10 or LOINC codes). The database also contains the patients' year of births and any date of deaths. |
| Columbia University Irving Medical Center (CUIMC) | The clinical data warehouse of NewYork-Presbyterian Hospital/Columbia University Irving Medical Center, New York, NY, based on its current and previous electronic health record systems, with data spanning over 30 years and including over 6 million patients |
| Information System for Research in Primary Care – Hospitalization Linked Data (SIDIAP-H) | The Information System for Research in Primary Care (SIDIAP; www.sidiap.org) is a primary care records database from Catalonia, North-East Spain. The SIDIAP-H subset of the database includes around 2 million people out of the total 7 million in SIDIAP that are registered in primary care |

| | |
|---|---|
| | practices with linked hospital inpatient data available as obtained from the Catalan Institute of Health hospitals. Healthcare is universal and tax-payer funded in the region, and primary care physicians are gatekeepers for all care and responsible for repeat prescriptions. |
| The Integrated Primary Care Information (IPCI) database | The Integrated Primary Care Information (IPCI) database is a longitudinal observational database containing electronics medical records from a representative sample (n=750) of general practitioners (GPs) in the Netherlands. The database contains up to 10 years of observational data from 2.36 Million persons. The IPCI data is collected from 9 different GP systems that are normalized into a common data structure. |

**Table S2**. Phenotype definitions: outcomes, well visit, chronic condition visit

| **Phenotype name** | **Adverse event Target Cohort ID (name and clean period)** | **Link to ATLAS cohort** | **Link to package SQL** |
|---|---|---|---|
| Acute myocardial infarction | 312: Acute myocardial infarction IP (365d) | https://atlas.ohdsi.org/#/cohortdefinition/340 | https://github.com/ohdsi-studies/Covid19VaccineAesiIncidenceCharacterization/blob/master/inst/sql/sql_server/outcome/AMI_IP.sql |
| Anaphylaxis | 303: Anaphylaxis (30d) | https://atlas.ohdsi.org/#/cohortdefinition/349 | https://github.com/ohdsi-studies/Covid19VaccineAesiIncidenceCharacterization/blob/master/inst/sql/sql_server/outcome/Anaphylaxis.sql |
| Appendicitis | 323: Appendicitis (365d) | https://atlas.ohdsi.org/#/cohortdefinition/386 | https://github.com/ohdsi-studies/Covid19VaccineAesiIncidenceCharacterization/blob/master/inst/sql/sql_server/outcome/Appendicitis.sql |
| Bell’s palsy | 307: Bells palsy (183d) | https://atlas.ohdsi.org/#/cohortdefinition/347 | https://github.com/ohdsi-studies/Covid19VaccineAesiIncidenceCharacterization/blob/master/inst/sql/sql_server/outcome/BellsPalsy.sql |
| Deep vein thrombosis | 332: Deep vein thrombosis cover ICD (365d) | https://atlas.ohdsi.org/#/cohortdefinition/402 | https://github.com/ohdsi-studies/Covid19VaccineAesiIncidenceCharacterization/blob/master/inst/sql/sql_server/outcome/DVT_coverICD.sql |

| | | | |
|---|---|---|---|
| Disseminated intravascular coagulation | 321: Disseminated intravascular coagulation (365d) | https://atlas.ohdsi.org/#/cohortdefinition/385 | https://github.com/ohdsi-studies/Covid19VaccineAesiIncidenceCharacterization/blob/master/inst/sql/sql_server/outcome/DisIntraCoag.sql |
| Encephalomyelitis | 308: Encephalomyelitis IP (183d) | https://atlas.ohdsi.org/#/cohortdefinition/346 | https://github.com/ohdsi-studies/Covid19VaccineAesiIncidenceCharacterization/blob/master/inst/sql/sql_server/outcome/Encephalomyelitis%20IP.sql |
| Guillain-Barre syndrome | 305: Guillain-Barre syndrome IP (365d) | https://atlas.ohdsi.org/#/cohortdefinition/343 | https://github.com/ohdsi-studies/Covid19VaccineAesiIncidenceCharacterization/blob/master/inst/sql/sql_server/outcome/GuillainBarreSyndrome%20IP.sql |
| Hemorrhagic stroke | 324: Hemorrhagic stroke IP (365d) | https://atlas.ohdsi.org/#/cohortdefinition/405 | https://github.com/ohdsi-studies/Covid19VaccineAesiIncidenceCharacterization/blob/master/inst/sql/sql_server/outcome/HemorrhagicStroke%20IP.sql |
| Immune thrombocytopenia | 326: Immune thrombocytopenia (365d) | https://atlas.ohdsi.org/#/cohortdefinition/335 | https://github.com/ohdsi-studies/Covid19VaccineAesiIncidenceCharacterization/blob/master/inst/sql/sql_server/outcome/ImmuneThrombocytopenia.sql |
| Myocarditis pericarditis | 314: Myocarditis pericarditis (365d) | https://atlas.ohdsi.org/#/cohortdefinition/339 | https://github.com/ohdsi-studies/Covid19VaccineAesiIncidenceCharacterization/blob/master/inst/sql/sql_server/outcome/MyocarditisPericarditis.sql |
| Narcolepsy | 319: Narcolepsy (365d) | https://atlas.ohdsi.org/#/cohortdefinition/345 | https://github.com/ohdsi-studies/Covid19VaccineAesiIncidenceCharacterization/blob/master/inst/sql/sql_server/outcome/Narcolepsy.sql |
| Non-hemorrhagic stroke | 310: Non-hemorrhagic stroke IP (365d) | https://atlas.ohdsi.org/#/cohortdefinition/406 | https://github.com/ohdsi-studies/Covid19VaccineAesiIncidenceCharacterization/blob/master/inst/sql/sql_server/outcome/NonHemorrhagicStroke%20IP.sql |
| Pulmonary embolism | 316: Pulmonary embolism (365d) | https://atlas.ohdsi.org/#/cohortdefinition/411 | https://github.com/ohdsi-studies/Covid19VaccineAesiIncidenceCharacterization/blob/master/inst/sql/sql_server/outcome/PulmonaryEmbolism.sql |

| | | | |
|---|---|---|---|
| Transverse myelitis | 318: Transverse myelitis (365d) | https://atlas.ohdsi.org/#/cohortdefinition/381 | https://github.com/ohdsi-studies/Covid19VaccineAesiIncidenceCharacterization/blob/master/inst/sql/sql_server/outcome/TransverseMyelitis.sql |
| Patients with a well visit | 110: persons at risk at start of year 2017-2019 with >=365d prior observation with >=1 well visit procedure in next year index 1JAN | | https://github.com/ohdsi-studies/Covid19VaccineAesiIncidenceCharacterization/blob/master/inst/sql/sql_server/target/1JAN_2017-19_w_well_visit.sql |
| Patients with chronic conditions | 105: persons at risk at start of year 2017-2019 with >=365d prior observation AND >=1 at risk condition excluding pregnancy | | https://github.com/ohdsi-studies/Covid19VaccineAesiIncidenceCharacterization/blob/master/inst/sql/sql_server/target/1JAN%202017-2019%20at-risk.sql |
| Patients with influenza vaccination | 107: persons at risk at start of year 2017-2019 with >=365d prior observation AND >=1 prior influenza vaccine | | https://github.com/ohdsi-studies/Covid19VaccineAesiIncidenceCharacterization/blob/master/inst/sql/sql_server/target/1JAN_2017-19_prior_flu_vax.sql |
| Pregnancy episodes | 126: persons at risk at start of year 2017-2019 with >=365d prior observation AND pregnant episode covering index (pregnancy start < 1JAN and pregnancy end >=1JAN) | | https://github.com/ohdsi-studies/Covid19VaccineAesiIncidenceCharacterization/blob/master/inst/sql/sql_server/target/1JAN%202017-2019%20pregnant.sql |

**Table S3.** Pooled age-adjusted incidence rate ratios for race and sex comparison, from meta-analyses, IRR and 95% CI.

| Outcome | Male versus female | Patients with race=Black versus patients with race=White |
|---|---|---|
| Acute myocardial infarction | 2.17 (1.89-2.49) | 1.12 (0.84-1.47) |
| Anaphylaxis | 0.87 (0.81-0.93) | 1.02 (0.91-1.15) |
| Appendicitis | 1.09 (1.02-1.15) | 0.67 (0.51-0.88) |
| Bell’s palsy | 1.05 (0.97-1.13) | 0.98 (0.79-1.22) |
| Deep vein thrombosis | 0.93 (0.86-1.02) | 1.13 (0.91-1.41) |
| Disseminated intravascular coagulation | 1.17 (0.99-1.38) | 1.49 (1.2-1.84) |
| Encephalomyelitis | 1.21 (1.11-1.31) | 1.25 (1.02-1.53) |
| Guillain-Barre syndrome | 1.37 (1.2-1.58) | 0.76 (0.6-0.96) |
| Hemorrhagic stroke | 1.39 (1.31-1.49) | 1.13 (0.94-1.35) |
| Immune thrombocytopenia | 0.96 (0.85-1.09) | 0.86 (0.7-1.07) |
| Myocarditis and pericarditis | 1.57 (1.45-1.69) | 1.25 (1.19-1.32) |
| Narcolepsy | 0.99 (0.85-1.14) | 0.81 (0.55-1.2) |
| Non-hemorrhagic stroke | 1.34 (1.25-1.45) | 1.48 (1.17-1.87) |
| Pulmonary embolism | 1.05 (1.01-1.1) | 1.29 (1.08-1.55) |
| Transverse myelitis | 0.76 (0.68-0.86) | 0.99 (0.9-1.1) |
| all | 1.15 (1.03-1.29) | 1.07 (0.97-1.18) |

**Figure S1.** Pooled age-adjusted incidence rate ratios (incidence rates for male versus female patients) from meta-analyses, IRR and 95% CI.

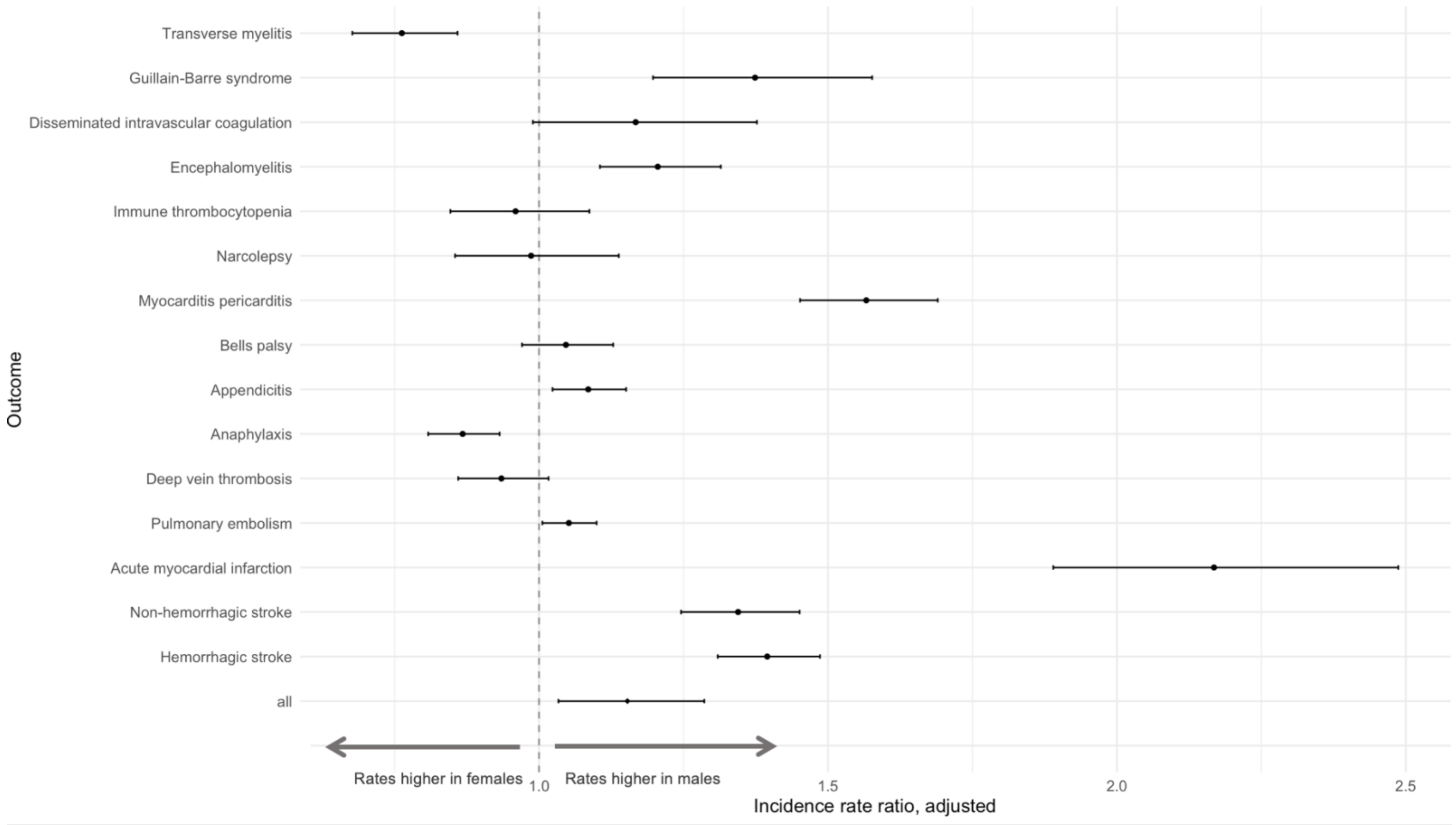

**Figure S2.** Pooled age-adjusted incidence rate ratios (incidence rates for patients with race=Black versus white patients with race=White) from meta-analyses, IRR and 95% CI.

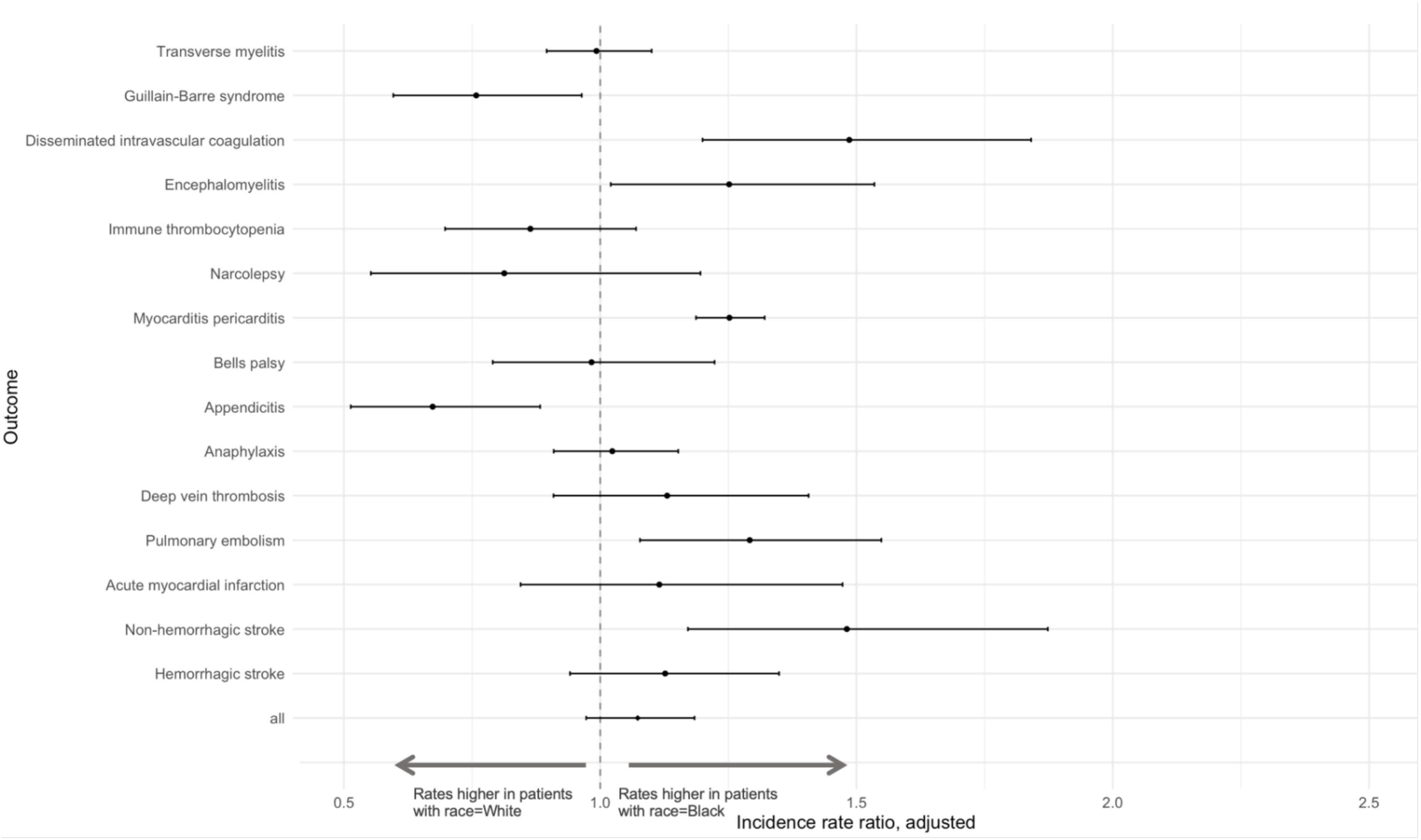

**Table S4.** Pooled age-adjusted incidence rate ratios (incidence rates of outcomes when entering the cohort on a visit versus entering on January 1st in patients with a visit in the next year) from meta-analyses, IRR and 95% CI.

| Outcome | Time at risk 0-1 days | Time at risk 1-28 days | Time at risk 1-42 days | Time at risk 1-90 days | Time at risk 1-365 days | Time at risk 0-365 days |
|---|---|---|---|---|---|---|
| Acute myocardial infarction | 24.72 (14.35-42.6) | 1.23 (1.12-1.35) | 1.08 (0.99-1.18) | 0.9 (0.82-1) | 0.83 (0.76-0.9) | 0.98 (0.95-1.01) |
| Anaphylaxis | 49.94 (33.83-73.73) | 1.48 (1.24-1.77) | 1.37 (1.18-1.58) | 1.13 (1.02-1.24) | 0.91 (0.88-0.94) | 1.09 (1.03-1.16) |
| Appendicitis | 51.79 (42.26-63.47) | 1.17 (1.01-1.36) | 1.01 (0.9-1.14) | 0.83 (0.77-0.9) | 0.79 (0.74-0.83) | 1.03 (0.99-1.07) |
| Bell's palsy | 34.17 (25.9-45.07) | 1.19 (0.95-1.49) | 1.05 (0.88-1.27) | 0.9 (0.79-1.01) | 0.86 (0.83-0.89) | 1.03 (0.98-1.09) |
| Deep vein thrombosis | 31.13 (22.48-43.1) | 1.42 (1.25-1.61) | 1.28 (1.16-1.42) | 1.09 (1.02-1.16) | 0.91 (0.89-0.93) | 1.03 (1-1.07) |
| Disseminated intravascular coagulation | 24.51 (15.31-39.23) | 2.04 (1.7-2.45) | 1.81 (1.53-2.14) | 1.44 (1.26-1.64) | 1.02 (0.94-1.09) | 1.13 (1.07-1.19) |
| Encephalomyelitis | 16.17 (10.09-25.89) | 1.92 (1.72-2.14) | 1.59 (1.47-1.71) | 1.24 (1.15-1.33) | 0.93 (0.88-0.97) | 1 (0.98-1.03) |
| Guillain-Barre syndrome | 20.77 (15.14-28.49) | 1.71 (1.46-2) | 1.43 (1.23-1.65) | 1.08 (0.98-1.19) | 0.87 (0.82-0.93) | 0.99 (0.95-1.02) |
| Hemorrhagic stroke | 27.14 (17.21-42.8) | 1.51 (1.38-1.66) | 1.3 (1.18-1.42) | 1.03 (0.93-1.14) | 0.86 (0.78-0.94) | 1.01 (0.98-1.04) |
| Immune thrombocytopenia | 25.39 (17.57-36.7) | 1.49 (1.26-1.77) | 1.33 (1.17-1.52) | 1.04 (0.96-1.13) | 0.89 (0.86-0.92) | 1 (0.97-1.04) |
| Myocarditis and pericarditis | 26.79 (17.04-42.13) | 1.47 (1.25-1.73) | 1.32 (1.16-1.5) | 1.03 (0.93-1.13) | 0.9 (0.86-0.93) | 1.03 (0.98-1.08) |
| Narcolepsy | 33.25 (22.52-49.07) | 1.15 (1.04-1.27) | 1.06 (0.98-1.14) | 0.96 (0.92-1.01) | 0.88 (0.85-0.91) | 1 (0.96-1.03) |
| Non-hemorrhagic stroke | 24.01 (13.42-42.94) | 1.34 (1.25-1.42) | 1.18 (1.12-1.24) | 0.96 (0.91-1.02) | 0.84 (0.79-0.9) | 0.98 (0.96-1.01) |
| Pulmonary embolism | 24.33 (17.3-34.22) | 1.41 (1.25-1.59) | 1.28 (1.16-1.41) | 1.06 (1-1.13) | 0.91 (0.89-0.94) | 1.03 (1.01-1.06) |
| Transverse myelitis | 17.33 (10-30.02) | 1.54 (1.25-1.89) | 1.36 (1.17-1.58) | 1.03 (0.92-1.16) | 0.88 (0.86-0.91) | 0.97 (0.94-1.01) |
| all | 28.04 (23.11-34.03) | 1.45 (1.33-1.57) | 1.28 (1.18-1.38) | 1.04 (0.98-1.1) | 0.89 (0.87-0.91) | 1.02 (1-1.03) |

**Figure S3.** Incidence rate ratio of incidence rate of outcomes when entering the cohort on a random visit versus entering on January 1st in patients with a visit in the next year, time-at-risk 0-1 day, 1-28 days, 1-42 days, 1-90 days and 1-365 days time-at-risk.

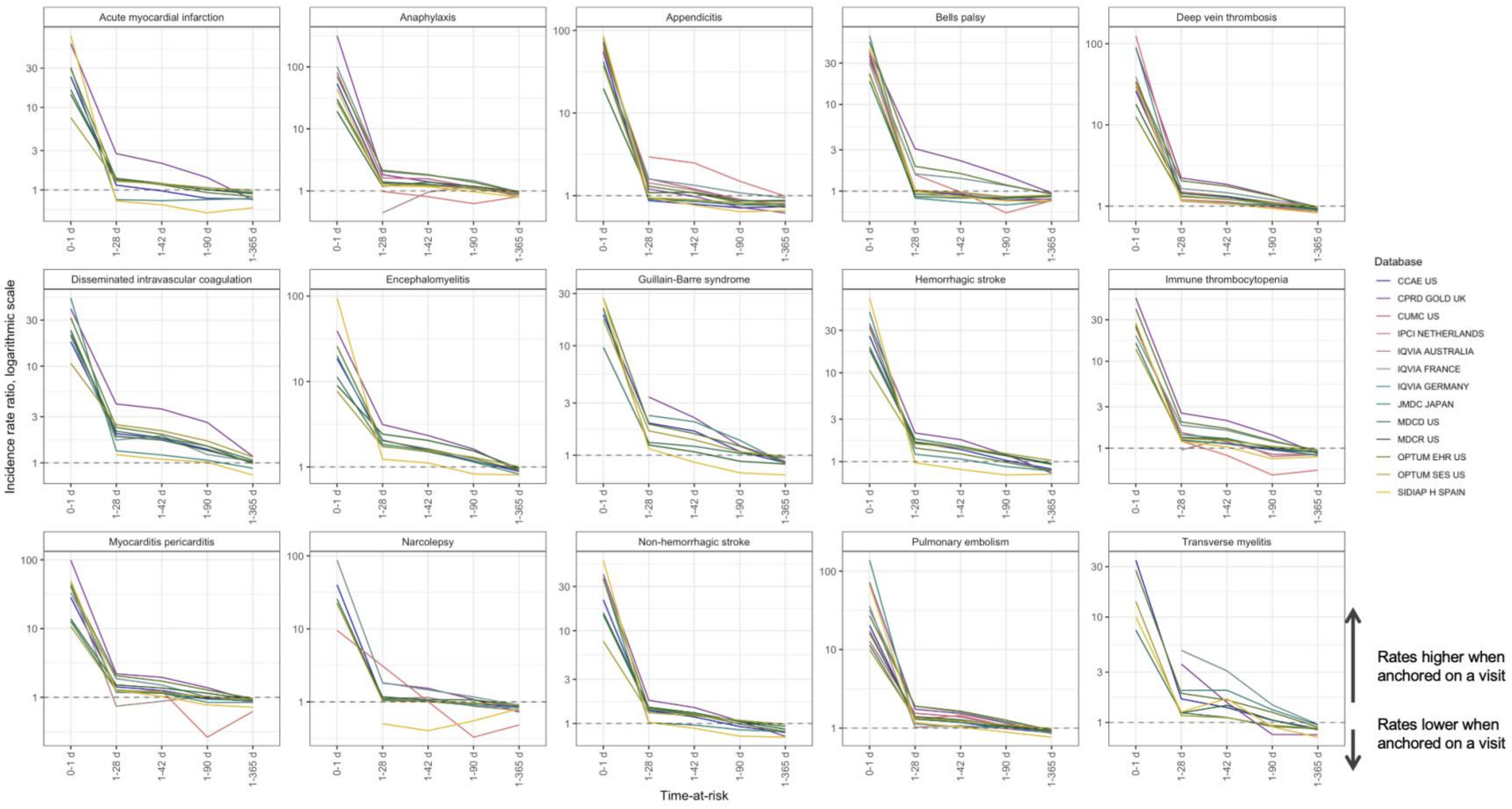

**Table S5.** Pooled age-adjusted incidence rate ratios (incidence rates of outcomes when entering the cohort on an influenza vaccination versus on January 1st with an influenza vaccination in the next year, from meta-analyses, IRR and 95% CI.

| Outcome | Time at risk 0-1 day | Time at risk 1-28 days | Time at risk 1-42 days | Time at risk 1-90 days | Time at risk 1-365 days |
|---|---|---|---|---|---|
| Acute myocardial infarction | 3.91 (1.46-10.46) | 1.28 (1.09-1.51) | 1.22 (1.06-1.4) | 1.15 (1.01-1.31) | 0.97 (0.82-1.13) |
| Anaphylaxis | 13.61 (8.97-20.65) | 1.12 (1.05-1.2) | 1.12 (1.06-1.19) | 1.05 (1.01-1.09) | 0.94 (0.92-0.96) |
| Appendicitis | 11.65 (6.81-19.94) | 0.9 (0.83-0.99) | 0.9 (0.83-0.97) | 0.85 (0.78-0.94) | 0.87 (0.81-0.94) |
| Bell's palsy | 8.84 (5.4-14.49) | 1.18 (0.97-1.44) | 1.12 (0.97-1.3) | 1.01 (0.92-1.11) | 0.95 (0.92-0.99) |
| Deep vein thrombosis | 12.19 (8.13-18.29) | 1.21 (1.07-1.36) | 1.19 (1.09-1.3) | 1.09 (1.04-1.15) | 0.95 (0.89-1.01) |
| Disseminated intravascular coagulation | 6.36 (3.62-11.18) | 1.79 (1.35-2.38) | 1.73 (1.36-2.2) | 1.71 (1.49-1.96) | 1.4 (1.23-1.59) |
| Encephalomyelitis | 2.02 (0.7-5.83) | 1.27 (0.96-1.67) | 1.22 (0.99-1.51) | 1.23 (1.03-1.47) | 1.04 (0.89-1.21) |
| Guillain-Barre syndrome | 5.18 (1.85-14.47) | 2.82 (2.07-3.86) | 2.2 (1.59-3.04) | 1.99 (1.54-2.56) | 1.4 (1.17-1.66) |
| Hemorrhagic stroke | 4.5 (1.52-13.29) | 1.38 (1.16-1.64) | 1.32 (1.11-1.57) | 1.26 (1.05-1.52) | 1.04 (0.86-1.27) |
| Immune thrombocytopenia | 16.21 (7.73-33.99) | 1.19 (1-1.43) | 1.14 (0.99-1.31) | 1.06 (0.96-1.18) | 0.92 (0.84-1) |
| Myocarditis and pericarditis | 7.11 (3.54-14.3) | 1.13 (0.93-1.38) | 1.1 (0.92-1.32) | 1.02 (0.94-1.11) | 0.94 (0.9-0.99) |
| Narcolepsy | 18.51 (9.64-35.53) | 1.05 (0.94-1.16) | 1 (0.9-1.11) | 0.97 (0.91-1.04) | 0.92 (0.9-0.95) |
| Non-hemorrhagic stroke | 4.17 (1.6-10.85) | 1.2 (1.07-1.35) | 1.17 (1.05-1.3) | 1.1 (0.98-1.23) | 0.95 (0.81-1.11) |
| Pulmonary embolism | 10.04 (6.56-15.36) | 1.11 (1.03-1.2) | 1.09 (1.03-1.16) | 1.05 (0.99-1.11) | 0.93 (0.84-1.04) |
| Transverse myelitis | 8.7 (2.75-27.48) | 1.08 (0.86-1.35) | 1.05 (0.87-1.25) | 0.98 (0.86-1.12) | 0.93 (0.87-1) |
| all | 8.68 (6.74-11.17) | 1.21 (1.12-1.32) | 1.16 (1.08-1.24) | 1.11 (1.04-1.18) | 0.97 (0.94-1.01) |

**Figure S4.** Incidence rate ratio of incidence rate of outcomes when entering the cohort on an influenza vaccination versus entering on January 1st in patients with an influenza vaccination in the next year, time-at-risk 1-28 days, 1-42 days, 1-90 days and 1-365 days time-at-risk.

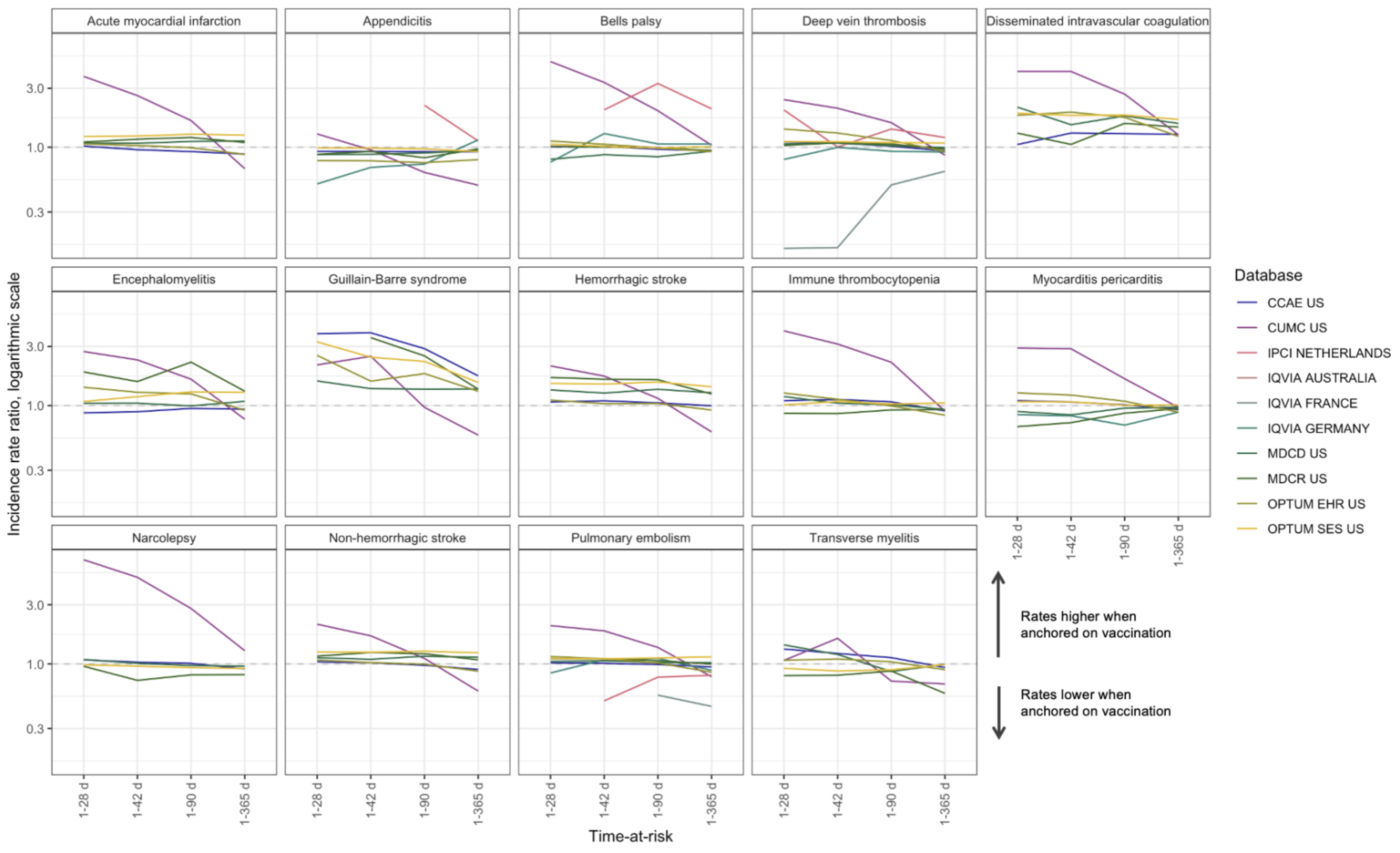

**Table S6.** Pooled age-adjusted incidence rate ratios (incidence rate of outcomes when entering the cohort on a well visit versus on January 1st in patients with a well visit in the next year) from meta-analyses, IRR and 95% CI.

| Outcome | Time at risk 0-1 day | Time at risk 1-28 days | Time at risk 1-42 days | Time at risk 1-90 days | Time at risk 1-365 days |
|---|---|---|---|---|---|
| Acute myocardial infarction | 1.14 (0.83-1.58) | 1.24 (1.19-1.29) | 1.19 (1.12-1.26) | 1.18 (1.12-1.24) | 1.13 (1.07-1.19) |
| Anaphylaxis | 18.08 (9.47-34.5) | 1.37 (1.3-1.44) | 1.33 (1.27-1.38) | 1.2 (1.15-1.25) | 0.95 (0.92-0.97) |
| Appendicitis | 3.44 (1.54-7.7) | 1.11 (1.08-1.16) | 1.11 (1.06-1.16) | 1.06 (1.03-1.08) | 1.01 (0.98-1.04) |
| Bell's palsy | 16.71 (9.86-28.29) | 0.98 (0.94-1.03) | 0.96 (0.91-1.01) | 0.93 (0.9-0.97) | 0.93 (0.91-0.96) |
| Deep vein thrombosis | 15.5 (9.29-25.86) | 1.16 (1.13-1.19) | 1.13 (1.11-1.15) | 1.09 (1.07-1.12) | 1.01 (0.98-1.04) |
| Disseminated intravascular coagulation | 6.92 (3.19-14.97) | 1.61 (1.2-2.17) | 1.49 (1.18-1.86) | 1.58 (1.34-1.86) | 1.3 (1.23-1.38) |
| Encephalomyelitis | 1.55 (0.61-3.91) | 1.27 (1.05-1.52) | 1.31 (1.13-1.52) | 1.31 (1.16-1.48) | 1.14 (1.08-1.2) |
| Guillain-Barre syndrome | 2.68 (0.91-7.88) | 1.2 (0.93-1.54) | 1.18 (0.96-1.46) | 1.14 (0.99-1.32) | 1.13 (1.04-1.22) |
| Hemorrhagic stroke | 0.88 (0.57-1.37) | 1.23 (1.08-1.41) | 1.29 (1.11-1.5) | 1.32 (1.16-1.5) | 1.22 (1.15-1.3) |
| Immune thrombocytopenia | 30.81 (14.03-67.65) | 1.22 (1.14-1.31) | 1.16 (1.09-1.23) | 0.97 (0.91-1.03) | 0.86 (0.78-0.95) |
| Myocarditis and pericarditis | 10.03 (5.44-18.49) | 1.08 (1.01-1.17) | 1.05 (0.98-1.11) | 0.99 (0.95-1.04) | 1 (0.98-1.02) |
| Narcolepsy | 29.85 (16.28-54.72) | 1.09 (1-1.19) | 1.06 (0.97-1.15) | 1 (0.95-1.05) | 0.92 (0.88-0.96) |
| Non-hemorrhagic stroke | 1.2 (0.59-2.42) | 1.25 (1.15-1.37) | 1.27 (1.16-1.38) | 1.25 (1.15-1.36) | 1.13 (1.08-1.19) |
| Pulmonary embolism | 12.32 (7.35-20.65) | 1.07 (1.01-1.13) | 1.06 (1.02-1.1) | 1.06 (1.04-1.09) | 1.03 (0.98-1.1) |
| Transverse myelitis | 15.59 (6.81-35.66) | 1.05 (0.87-1.25) | 1.02 (0.88-1.18) | 1.05 (0.95-1.17) | 0.92 (0.87-0.97) |
| all | 6.32 (3.1-12.89) | 1.17 (1.11-1.23) | 1.15 (1.09-1.2) | 1.11 (1.06-1.16) | 1.04 (0.99-1.09) |

**Figure S5.** Incidence rate ratio of incidence rate of outcomes when entering the cohort on a well visit versus entering on January 1st in patients with a well visit in the next year, time-at-risk 1-28 days, 1-42 days, 1-90 days and 1-365 days time-at-risk.

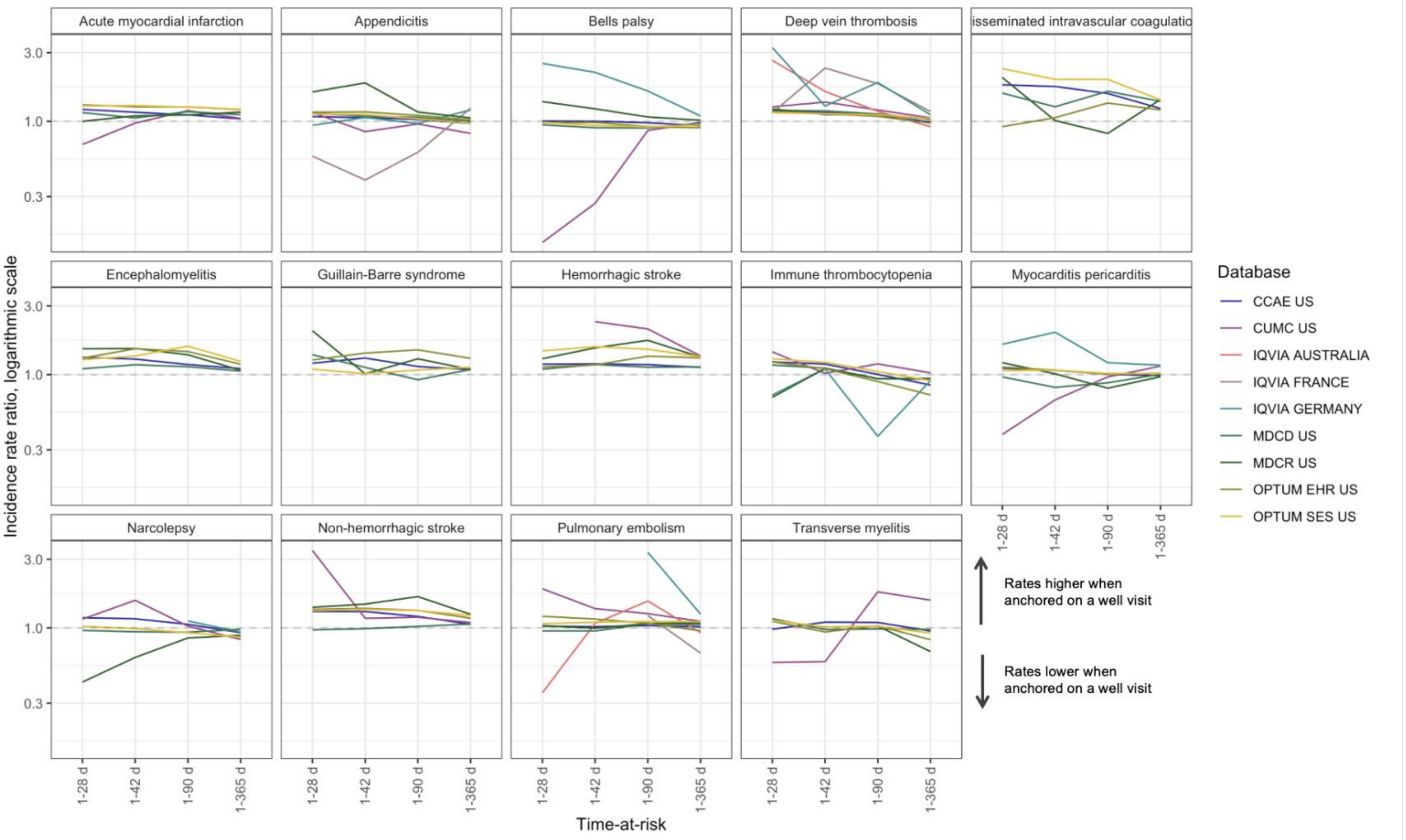

**Table S7.** Seasonal trends. Incidence rate ratios of incidence rates for quarter 2 (April - June), quarter 3 (July - September) and quarter 4 to quarter 1 (January – March) of 2017-2019 from meta-analysis, IRR and 95% CI.

| Outcome | Quarter 2 versus Quarter 1 | Quarter 3 versus Quarter 1 | Quarter 4 versus Quarter 1 |
|---|---|---|---|
| Acute myocardial infarction | 1.1 (1.01-1.21) | 0.95 (0.9-0.99) | 0.95 (0.9-0.99) |
| Anaphylaxis | 1.11 (1.02-1.2) | 1.32 (1.22-1.42) | 1.32 (1.22-1.42) |
| Appendicitis | 1.11 (1.02-1.21) | 1.04 (1-1.09) | 1.04 (1-1.09) |
| Bell's palsy | 1.09 (1.01-1.18) | 0.99 (0.95-1.03) | 0.99 (0.95-1.03) |
| Deep vein thrombosis | 1.07 (1-1.15) | 1.06 (1.03-1.09) | 1.06 (1.03-1.09) |
| Disseminated intravascular coagulation | 1.04 (0.96-1.12) | 0.93 (0.87-1) | 0.93 (0.87-1) |
| Encephalomyelitis | 1.13 (1-1.29) | 1.01 (0.94-1.09) | 1.01 (0.94-1.09) |
| Guillain-Barre syndrome | 1.08 (0.97-1.19) | 0.95 (0.86-1.04) | 0.95 (0.86-1.04) |
| Hemorrhagic stroke | 1.06 (1.01-1.11) | 0.99 (0.95-1.03) | 0.99 (0.95-1.03) |
| Immune thrombocytopenia | 1.08 (1-1.17) | 0.97 (0.93-1.01) | 0.97 (0.93-1.01) |
| Myocarditis and pericarditis | 1.09 (0.98-1.2) | 0.91 (0.84-0.98) | 0.91 (0.84-0.98) |
| Narcolepsy | 1.07 (1.02-1.12) | 1.04 (1.01-1.07) | 1.04 (1.01-1.07) |
| Non-hemorrhagic stroke | 1.07 (0.99-1.15) | 1 (0.96-1.03) | 1 (0.96-1.03) |
| Pulmonary embolism | 1.09 (1.01-1.17) | 0.98 (0.95-1.01) | 0.98 (0.95-1.01) |
| Transverse myelitis | 1.05 (0.92-1.19) | 0.97 (0.91-1.02) | 0.97 (0.91-1.02) |
| all | 1.08 (1.06-1.1) | 1 (0.97-1.03) | 1 (0.97-1.03) |

**Figure S6.** Comparison of incidence rates for quarter 2 (April - June), quarter 3 (July - September) and quarter 4 to quarter 1 (January – March) of 2017-2019 from meta-analysis, IRR and 95% CI.

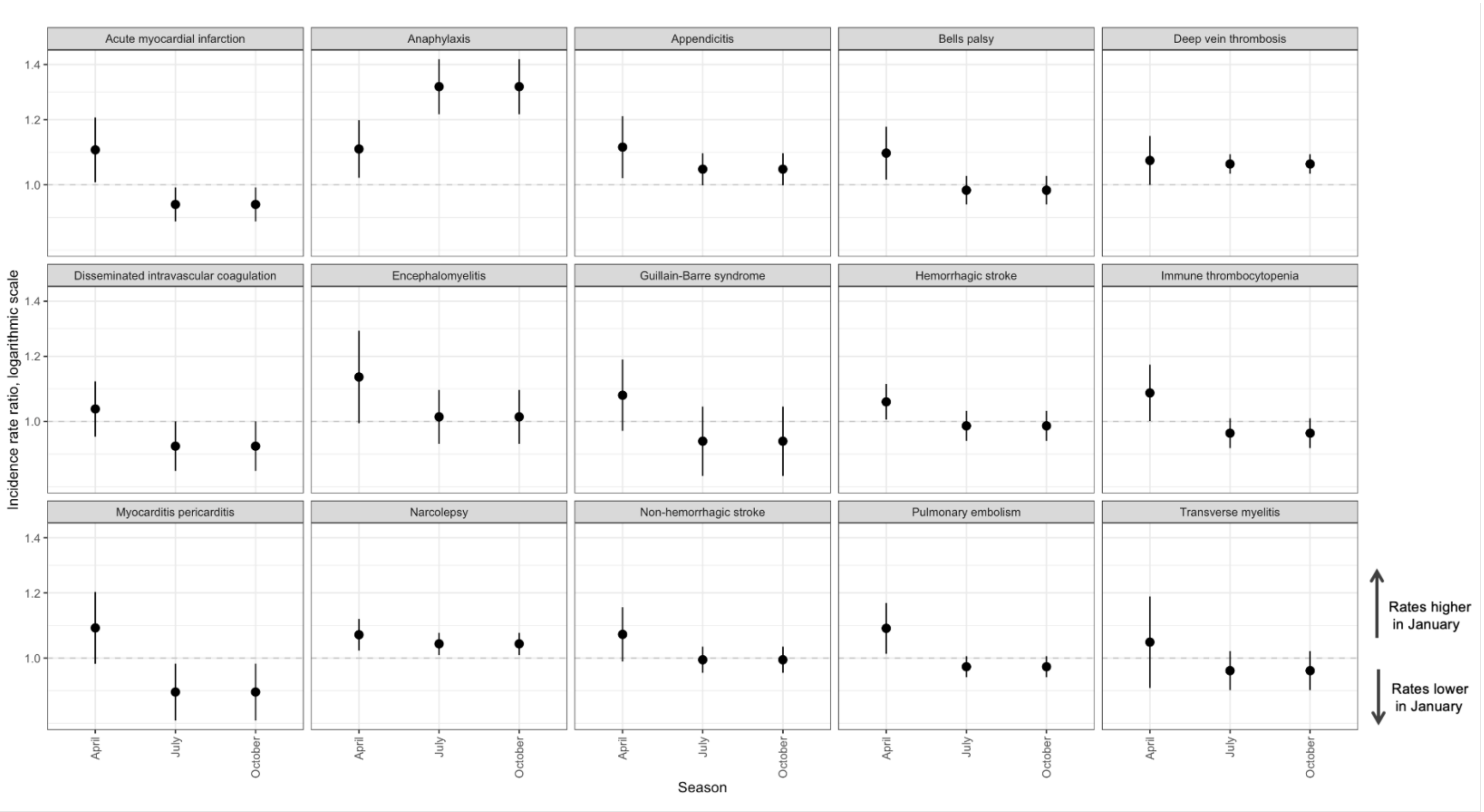

**Table S8.** Incidence rate ratios of incidence rates for Quarter 2 and 3 of 2020 versus Quarter 2 and 3 of 2017 – 2019 from meta-analysis, IRR and 95% CI.

| Outcome | Quarter 2 2020 versus Quarter 2 2017-2019 | Quarter 3 2020 versus Quarter 3 2017-2019 |
|---|---|---|
| Acute myocardial infarction | 0.95 (0.9-0.99) | 0.92 (0.77-1.09) |
| Anaphylaxis | 1.32 (1.22-1.42) | 1.28 (0.93-1.76) |
| Appendicitis | 1.04 (1-1.09) | 1.16 (0.91-1.49) |
| Bell’s palsy | 0.99 (0.95-1.03) | 1.12 (0.9-1.41) |
| Deep vein thrombosis | 1.06 (1.03-1.09) | 1.04 (0.86-1.24) |
| Disseminated intravascular coagulation | 0.93 (0.87-1) | 0.81 (0.63-1.04) |
| Encephalomyelitis | 1.01 (0.94-1.09) | 0.84 (0.63-1.11) |
| Guillain-Barre syndrome | 0.95 (0.86-1.04) | 0.83 (0.55-1.26) |
| Hemorrhagic stroke | 0.99 (0.95-1.03) | 0.85 (0.7-1.04) |
| Immune thrombocytopenia | 0.97 (0.93-1.01) | 1.08 (0.89-1.3) |
| Myocarditis and pericarditis | 0.91 (0.84-0.98) | 1.18 (0.86-1.62) |
| Narcolepsy | 1.04 (1.01-1.07) | 1.15 (0.86-1.52) |
| Non-hemorrhagic stroke | 1 (0.96-1.03) | 0.95 (0.77-1.17) |
| Pulmonary embolism | 0.98 (0.95-1.01) | 1.25 (1.03-1.52) |
| Transverse myelitis | 0.97 (0.91-1.02) | 1.09 (0.93-1.27) |
| all | 1 (0.97-1.03) | 1.03 (0.96-1.1) |

**Figure S7.** Comparison of incidence rates in Q2, Q3 of 2017 – 2019 (pre-COVID-19 pandemic) versus corresponding quarters in 2020 (COVID-19 pandemic), incidence rate ratios.

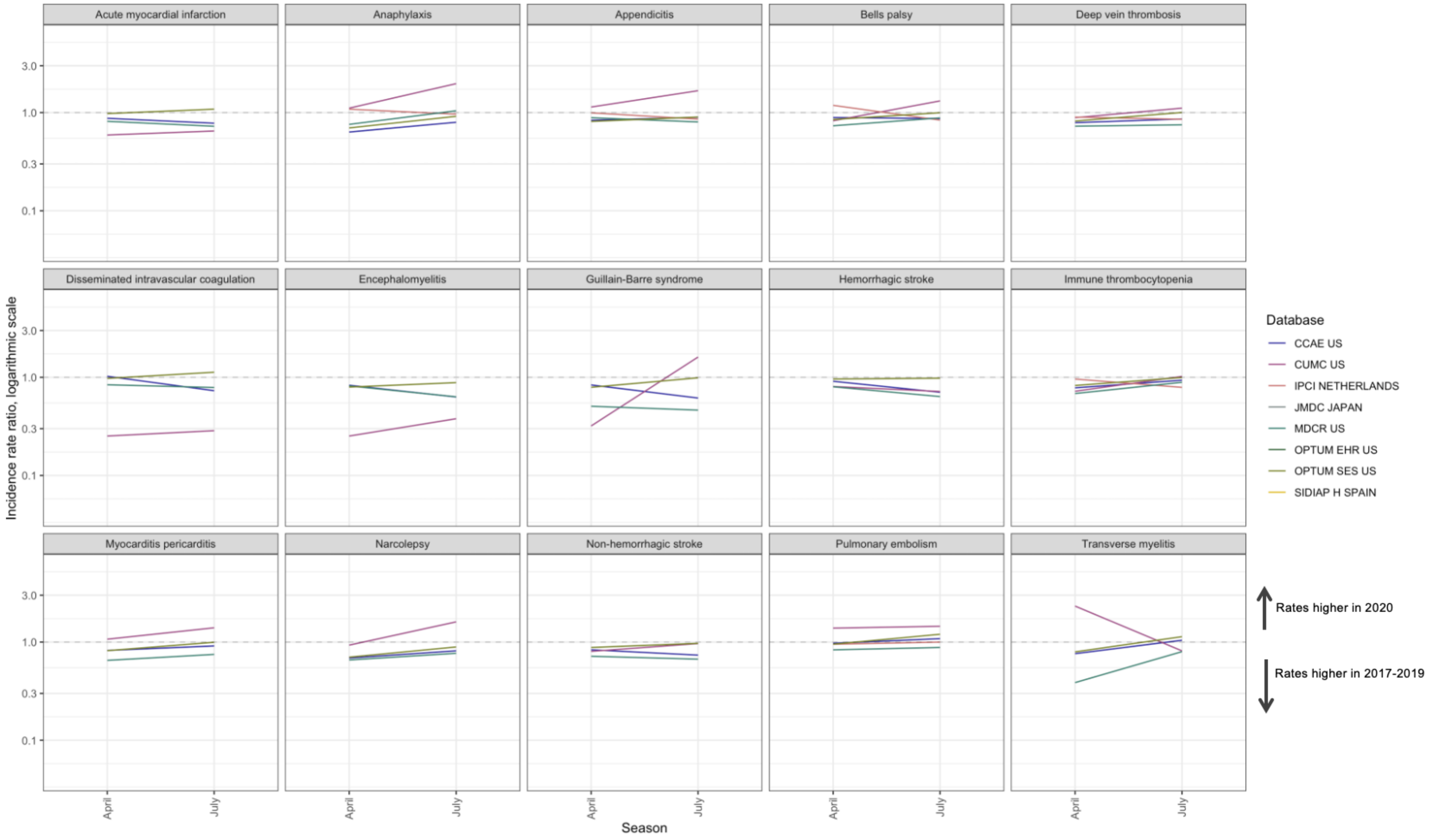

**Table S9.** Incidence rate ratios of incidence rates for patient subgroups from meta-analysis, IRR and 95% CI.

| Outcome | Patients with prior influenza vaccine versus general population | Patients with chronic conditions versus patients with no chronic conditions |
|---|---|---|
| Acute myocardial infarction | 1.48 (1.21-1.8) | 2.94 (2.29-3.79) |
| Anaphylaxis | 1.41 (1.26-1.59) | 1.78 (1.57-2.02) |
| Appendicitis | 1.09 (0.99-1.2) | 1.34 (1.2-1.49) |
| Bell’s palsy | 1.35 (1.17-1.55) | 2.08 (1.87-2.31) |
| Deep vein thrombosis | 1.49 (1.28-1.74) | 2.59 (2.25-2.98) |
| Disseminated intravascular coagulation | 1.69 (1.33-2.14) | 4.06 (2.95-5.59) |
| Encephalomyelitis | 1.61 (1.3-1.99) | 2.84 (2.33-3.47) |
| Guillain-Barre syndrome | 1.01 (0.84-1.22) | 2.03 (1.61-2.56) |
| Hemorrhagic stroke | 1.44 (1.19-1.75) | 2.19 (1.74-2.76) |
| Immune thrombocytopenia | 1.62 (1.41-1.85) | 2.35 (2.05-2.7) |
| Myocarditis and pericarditis | 1.99 (1.63-2.43) | 2.94 (2.44-3.54) |
| Narcolepsy | 1.21 (1.03-1.42) | 1.93 (1.63-2.29) |
| Non-hemorrhagic stroke | 1.47 (1.19-1.8) | 2.58 (2.01-3.29) |
| Pulmonary embolism | 1.54 (1.33-1.78) | 2.75 (2.35-3.22) |
| Transverse myelitis | 1.27 (1.08-1.49) | 1.65 (1.37-1.99) |
| all | 1.41 (1.29-1.55) | 2.29 (1.99-2.64) |

**Figure S8.** Comparison of incidence rates for patients with chronic conditions versus patients with no chronic conditions (A) and patients with prior influenza vaccination versus general population (B) from meta-analysis, IRR and 95% CI.

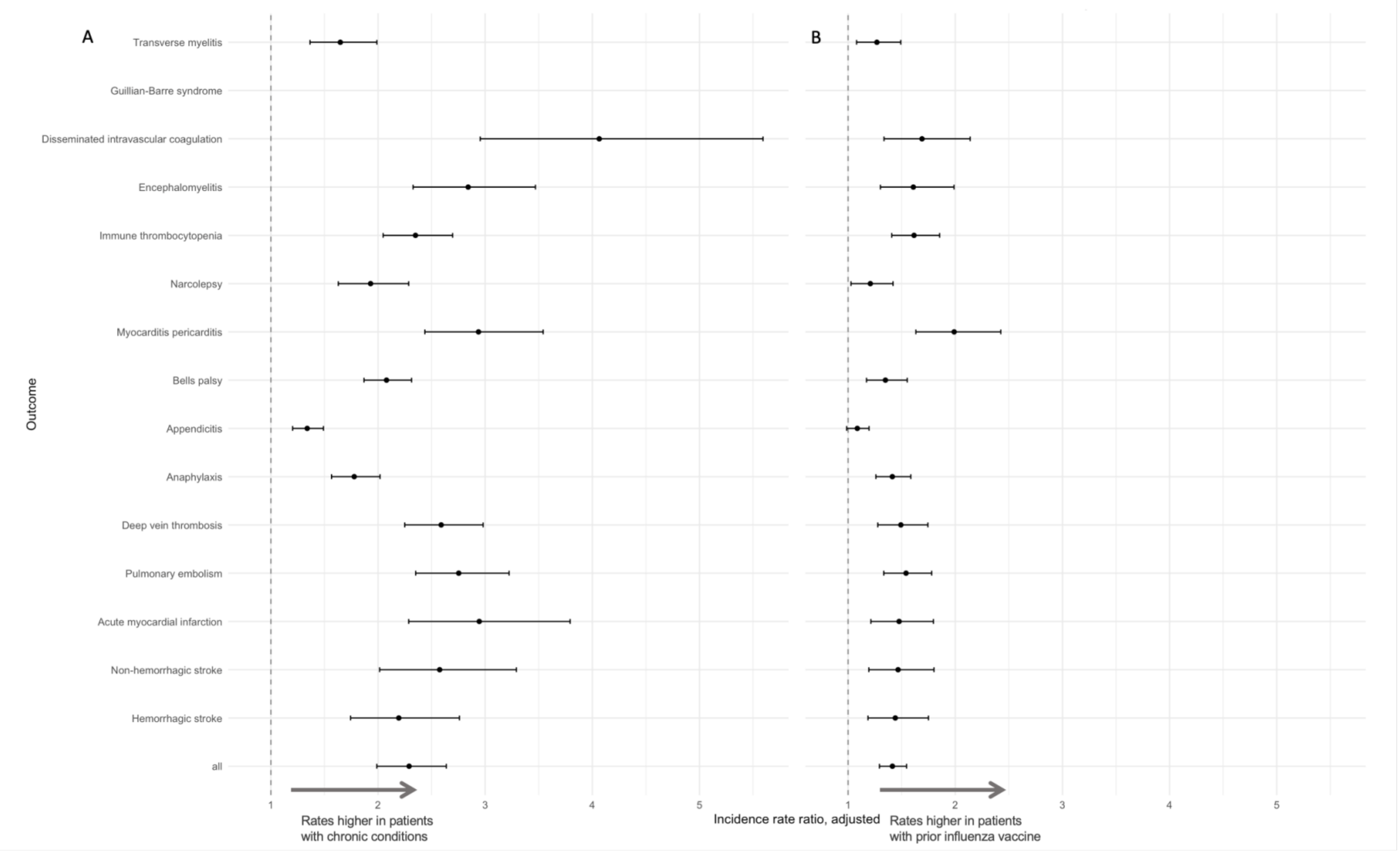

**Table S10.** Pooled age-adjusted incidence rate ratios for comparing clean windows and prior observation, from meta-analyses, IRR and 95% CI.

| Outcome | First ever conditions in patient history versus first occurrence in a given window | Requirement to have a year of prior observation versus no prior observation requirement |
|---|---|---|
| Acute myocardial infarction | 0.91 (0.89-0.93) | 1.01 (0.93-1.09) |
| Anaphylaxis | 0.7 (0.66-0.73) | 1.14 (1.04-1.26) |
| Appendicitis | 0.98 (0.96-0.99) | 0.99 (0.93-1.05) |
| Bell’s palsy | 0.77 (0.73-0.8) | 0.96 (0.89-1.05) |
| Deep vein thrombosis | 0.82 (0.8-0.84) | 0.96 (0.85-1.09) |
| Disseminated intravascular coagulation | 0.96 (0.95-0.98) | 0.77 (0.7-0.85) |
| Encephalomyelitis | 0.93 (0.91-0.95) | 0.99 (0.86-1.14) |
| Guillain-Barre syndrome | 0.59 (0.48-0.71) | 0.98 (0.85-1.14) |
| Hemorrhagic stroke | 0.96 (0.96-0.97) | 1 (0.92-1.1) |
| Immune thrombocytopenia | 0.7 (0.67-0.73) | 0.96 (0.78-1.19) |
| Myocarditis and pericarditis | 0.89 (0.86-0.91) | 0.95 (0.9-1) |
| Narcolepsy | 0.69 (0.65-0.74) | 0.8 (0.68-0.94) |
| Non-hemorrhagic stroke | 0.88 (0.86-0.91) | 1.04 (0.95-1.15) |
| Pulmonary embolism | 0.8 (0.78-0.82) | 0.89 (0.77-1.04) |
| Transverse myelitis | 0.71 (0.67-0.75) | 0.75 (0.62-0.91) |
| all | 0.82 (0.78-0.86) | 0.95 (0.9-1) |

**Figure S9.** Comparison of prior observation (A) and clean windows (B) , from meta-analyses, IRR and 95% CI.

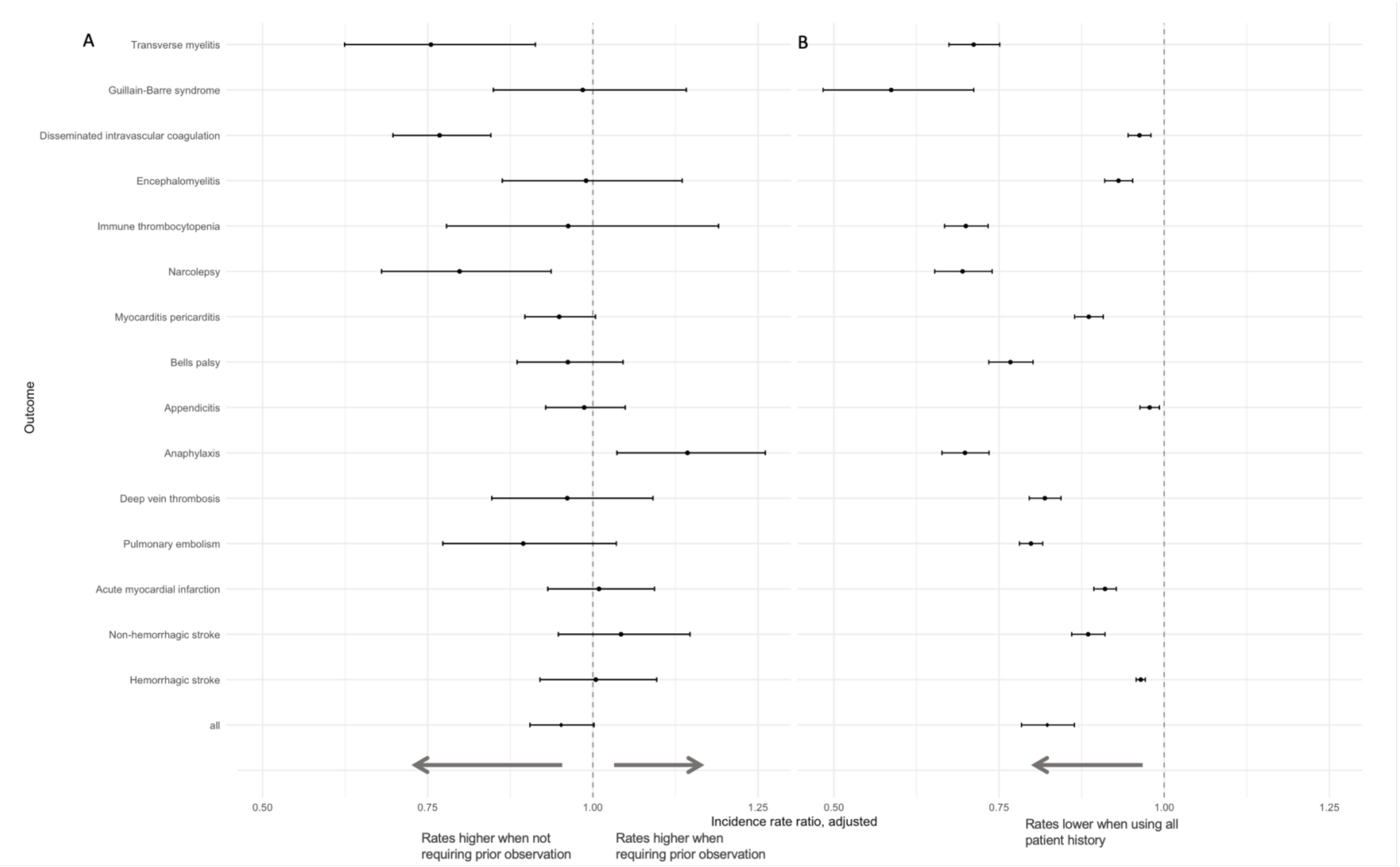

**Table S11.** Pooled estimated age-sex stratified incidence rates per 100,00 person-years (with 95% confidence intervals), calculated from meta-analyses.

| | | Age group | | | | | | | |
|---|---|---|---|---|---|---|---|---|---|
| **Outcome** | **Sex** | **1 - 5** | **6 - 17** | **18 - 34** | **35 - 54** | **55 - 64** | **65 - 74** | **75 - 84** | **85+** |
| Non-hemorrhagic stroke | Female | 4 (2-9) | 4 (1-12) | 18 (4-86) | 83 (11-617) | 217 (25-1882) | 413 (77-2198) | 874 (197-3884) | 1523 (320-7239) |
| | Male | 6 (2-20) | 5 (2-10) | 17 (4-75) | 119 (21-664) | 370 (67-2046) | 612 (145-2578) | 1063 (242-4662) | 1495 (260-8607) |
| Acute myocardial infarction | Female | <1 (<1-1) | <1 (<1-1) | 6 (1-49) | 54 (7-430) | 171 (24-1235) | 312 (76-1280) | 617 (184-2069) | 1144 (313-4184) |
| | Male | <1 (<1-1) | 1 (1-1) | 16 (4-72) | 172 (40-740) | 467 (135-1611) | 653 (214-1994) | 934 (290-3013) | 1514 (356-6432) |
| Deep vein thrombosis | Female | 12 (3-50) | 18 (8-40) | 140 (66-298) | 306 (117-797) | 428 (150-1224) | 683 (257-1820) | 975 (360-2642) | 1206 (407-3572) |
| | Male | 14 (4-55) | 14 (6-32) | 80 (28-228) | 272 (88-836) | 499 (194-1289) | 695 (250-1931) | 831 (254-2720) | 1003 (278-3616) |
| Hemorrhagic stroke | Female | 7 (2-28) | 5 (2-16) | 13 (4-47) | 36 (7-175) | 77 (15-389) | 124 (29-527) | 249 (56-1108) | 412 (85-1986) |
| | Male | 8 (2-43) | 8 (3-24) | 19 (5-76) | 51 (10-268) | 115 (23-562) | 178 (49-650) | 312 (73-1340) | 506 (86-2961) |
| Pulmonary embolism | Female | 1 (<1-36) | 3 (1-13) | 38 (11-124) | 81 (21-309) | 125 (33-470) | 217 (77-611) | 358 (135-951) | 427 (154-1184) |
| | Male | 1 (<1-24) | 2 (<1-12) | 20 (5-80) | 80 (20-318) | 171 (59-497) | 256 (96-683) | 349 (119-1030) | 398 (124-1277) |
| Appendicitis | Female | 32 (12-84) | 154 (55-430) | 134 (69-260) | 85 (42-172) | 66 (28-156) | 53 (20-143) | 40 (13-124) | 35 (12-98) |
| | Male | 38 (17-85) | 194 (101-372) | 146 (81-266) | 88 (49-159) | 65 (32-132) | 57 (23-144) | 47 (15-152) | 45 (14-143) |
| Bells palsy | Female | 15 (9-27) | 25 (12-51) | 44 (23-84) | 61 (26-140) | 76 (31-184) | 86 (29-256) | 101 (31-330) | 92 (31-274) |
| | Male | 15 (10-24) | 21 (13-34) | 43 (29-64) | 68 (37-125) | 86 (43-172) | 94 (35-252) | 92 (29-291) | 100 (34-292) |
| Anaphylaxis | Female | 49 (16-150) | 50 (16-154) | 39 (16-95) | 34 (13-91) | 35 (14-85) | 29 (11-76) | 23 (7-73) | 12 (4-36) |

| | | | | | | | | | |
|---|---|---|---|---|---|---|---|---|---|
| | Male | 74 (26-209) | 56 (18-175) | 29 (14-63) | 24 (11-53) | 25 (11-53) | 24 (9-68) | 18 (7-49) | 10 (2-50) |
| Immune thrombocytopenia | Female | 12 (8-19) | 9 (4-21) | 14 (6-36) | 15 (5-43) | 18 (6-53) | 25 (8-82) | 30 (8-110) | 36 (11-118) |
| | Male | 17 (12-23) | 8 (3-19) | 8 (2-23) | 10 (3-35) | 19 (6-57) | 30 (9-105) | 41 (10-170) | 56 (15-210) |
| Myocarditis pericarditis | Female | 6 (1-25) | 7 (2-21) | 16 (8-32) | 22 (9-53) | 31 (13-72) | 35 (12-97) | 39 (11-138) | 34 (8-143) |
| | Male | 7 (1-32) | 11 (5-24) | 37 (16-88) | 37 (16-87) | 45 (20-102) | 49 (17-139) | 54 (15-193) | 41 (9-193) |
| Disseminated intravascular coagulation | Female | 2 (<1-104) | 2 (<1-48) | 4 (<1-99) | 5 (<1-75) | 10 (1-89) | 14 (2-97) | 19 (4-94) | 16 (3-82) |
| | Male | 3 (<1-137) | 2 (<1-44) | 4 (<1-31) | 5 (1-56) | 12 (1-120) | 17 (2-154) | 23 (4-152) | 24 (5-126) |
| Encephalomyelitis | Female | 5 (2-15) | 5 (2-16) | 5 (2-19) | 6 (1-44) | 9 (1-61) | 11 (2-62) | 12 (2-77) | 14 (2-100) |
| | Male | 5 (2-12) | 5 (2-14) | 5 (2-17) | 7 (1-55) | 12 (3-58) | 16 (3-73) | 18 (3-101) | 16 (1-180) |
| Narcolepsy | Female | 1 (<1-5) | 7 (3-17) | 15 (4-52) | 11 (2-55) | 9 (2-42) | 10 (2-46) | 8 (1-49) | 9 (2-42) |
| | Male | 1 (<1-5) | 6 (2-18) | 13 (4-40) | 10 (2-47) | 11 (3-44) | 10 (2-50) | 10 (2-68) | 10 (2-60) |
| Guillain-Barre syndrome | Female | 1 (<1-8) | 1 (<1-2) | 3 (1-5) | 3 (1-11) | 5 (1-18) | 6 (2-19) | 6 (3-16) | 7 (2-22) |
| | Male | 2 (<1-18) | 1 (<1-3) | 2 (1-4) | 4 (2-7) | 7 (4-14) | 8 (3-25) | 11 (3-40) | 12 (2-68) |
| Transverse myelitis | Female | 1 (<1-3) | 1 (<1-3) | 3 (1-8) | 4 (1-12) | 4 (2-13) | 4 (2-13) | 4 (1-11) | 2 (1-9) |
| | Male | 1 (<1-2) | 1 (<1-3) | 2 (1-6) | 3 (1-10) | 4 (1-10) | 4 (1-11) | 4 (1-13) | 4 (1-11) |
